# A Randomized Trial Comparing Omicron-Containing Boosters with the Original Covid-19 Vaccine mRNA-1273

**DOI:** 10.1101/2023.01.24.23284869

**Authors:** Ivan T. Lee, Catherine A. Cosgrove, Patrick Moore, Claire Bethune, Rhiannon Nally, Marcin Bula, Philip A. Kalra, Rebecca Clark, Paul I. Dargan, Marta Boffito, Ray Sheridan, Ed Moran, Thomas C. Darton, Fiona Burns, Dinesh Saralaya, Christopher J. A. Duncan, Patrick Lillie, Alberto San Francisco Ramos, Eva Galiza, Paul T. Heath, Bethany Girard, Christy Parker, Dondi Rust, Shraddha Mehta, Elizabeth de Windt, Andrea Sutherland, Joanne E. Tomassini, Frank J. Dutko, Spyros Chalkias, Weiping Deng, Xing Chen, LaRee Tracy, Honghong Zhou, Jacqueline M. Miller, Rituparna Das, the Study Investigators

## Abstract

**Background:** Omicron-containing bivalent boosters are available worldwide. Results of a large, randomized, active-controlled study are presented.

**Methods:** This phase 3, randomized, observer-blind, active-controlled trial in the United Kingdom evaluated the immunogenicity and safety of 50-µg doses of omicron-BA.1-monovalent mRNA-1273.529 and bivalent mRNA-1273.214 booster vaccines compared with 50-µg mRNA-1273 administered as boosters in individuals ≥16 years. Participants had previously received 2 doses of any authorized/approved Covid-19 vaccine with or without an mRNA vaccine booster. Safety and immunogenicity were primary objectives; immunogenicity was assessed in all participants, with analysis conducted based on prior infection status. Incidence of Covid-19 post-boost was a secondary (mRNA-1273.214) or exploratory (mRNA-1273.529) objective.

**Results:** In part 1 of the study, 719 participants received mRNA-1273.529 (n=362) or mRNA-1273 (n=357); in part 2, 2813 received mRNA-1273.214 (n=1418) or mRNA-1273 (n=1395). Median durations (months [interquartile range]) between the most recent Covid-19 vaccine and study boosters were similar in the mRNA-1273.529 (4.0 [3.6-4.7]) and mRNA-1273 (4.1 [3.5-4.7]) (part 1), and mRNA-1273.214 (5.5 [4.8-6.2] and mRNA-1273 (5.4 [4.8-6.2]) groups (part 2).

Both mRNA-1273.529 and mRNA-1273.214 elicited superior neutralizing antibody responses against omicron BA.1 with geometric mean ratios (99% CIs) of 1.68 (1.45-1.95) and 1.53 (1.41-1.67) compared to mRNA-1273 at day 29 post-boost. Although the study was not powered to assess relative vaccine efficacy, the incidence rates/1000 person years (95% CI) of Covid-19 trended lower with mRNA-1273.529 (670.5 [528.3-839.3]) than mRNA-1273 (769.3 [615.4-950.1]) and mRNA-1273.214 (633.0 [538.1-739.7]) than mRNA-1273 (711.6 [607.5-828.5]).

Sequence analysis in part 2 showed that this was driven by lower incidence of Covid-19 in the mRNA-1273.214 cohort with BA.2 and BA.4 sublineages but not BA.5 sublineages. All study boosters were well-tolerated.

**Conclusion:** The bivalent omicron BA.1-containing booster elicited superior neutralizing antibody responses against omicron BA.1 with acceptable safety results consistent with the BA.1 monovalent vaccine. Incidence rates for Covid-19 were numerically lower in participants who received mRNA-1273.214 compared to the original booster vaccine mRNA-1273, driven by the BA.2 and BA.4 sublineages.

## Introduction

The continuous and rapid evolution of severe acute respiratory syndrome coronavirus 2 (SARS-CoV-2) and the emergence of viral variants with increased transmissibility and antibody escape from infection- or vaccine-induced immunity has required booster immunization to maintain protection against coronavirus disease 2019 (Covid-19). After the emergence of omicron sublineages, variant-targeting bivalent booster vaccines comprised of mRNAs encoding for the original SARS-CoV-2 and for omicron BA.1 or BA.4/BA.5 have been deployed to prevent Covid-19 caused by divergent variants.^1-5^ Variant evolution has since continued along the omicron lineages with new sublineages BQ1.1, XBB.1 and XBB1.5 rising in different geographies. This has raised questions about the timing and composition of additional vaccine updates should they be deemed necessary.^4,6^

In prior, non-randomized clinical studies, the bivalent omicron BA.1-containing mRNA-1273.214 and omicron-BA.4/BA.5-containing mRNA-1273.222 booster vaccines elicited superior neutralizing antibody responses against omicron BA.1 and BA.4/BA.5, respectively, versus mRNA-1273 with no new safety concerns.^1-3^ Recent real-world evidence has shown that the BA.1 and BA.4/5 containing bivalent boosters provided additional protection against Covid-19 compared to those immunized with the original vaccine boosters or unvaccinated individuals.^7-9^ However, effectiveness studies of the contemporaneous administration of the bivalent or original vaccine have not been undertaken.

Safety and immunogenicity data from open-label studies, non-randomized, non-contemporaneously controlled studies have supported the authorization of mRNA-1273 omicron-targeting bivalent booster vaccines in rapid response to variant surges.^1-3^ The mRNA-BA.1-containing mRNA-1273.214 booster was approved in the United Kingdom (UK) by the Medicines and Healthcare products Regulatory Agency in August 2022 and as the predominance of omicron variants transitioned from omicron BA.1 and sublineages to BA.4/BA.5 lineages, the mRNA-1273.222 booster was subsequently approved.^5,10-13^ Here, we describe interim results from a large, phase 3 randomized, observer-blind, active-controlled clinical trial that evaluated the safety and immunogenicity of 50 µg of omicron-BA.1 containing bivalent mRNA-1273.214 (25 µg ancestral SARS-CoV-2 and 25 µg of omicron BA.1 spike mRNA) and 50 µg monovalent mRNA-1273.529 (contains only omicron BA.1 spike mRNA) booster vaccines compared with 50 µg of the original mRNA-1273 booster in individuals aged ≥16 years in the UK. We also summarize interim data on the incidence rates of Covid-19 post-booster.

## Methods

### Trial Design and Participants

This large, phase 2/3 (designated as phase 3 having >3000 participants)^14^ two-part, randomized, observer-blind, active-controlled, multicenter (28 sites) trial evaluated the immunogenicity and safety of 50-µg mRNA-1273.529 (part 1) and 50-µg mRNA-1273.214 (part 2) booster vaccines compared with 50-µg mRNA-1273 in medically-stable individuals aged ≥16 years in the United Kingdom (UK; EudraCT, 2022-000063-51) (Fig. S1). The trial was initiated in February of 2022 with a BA.1 monovalent booster vaccine in response to the emergence of the omicron variant.

Thereafter, due to the rapid evolution of omicron sublineages, the enrollment of the BA.1 monovalent portion of the trial (part 1) was stopped to expedite enrollment of a BA.1 bivalent vaccine (part 2), which was hypothesized to induce better cross-protection.

Eligible participants had previously received 2 injections of an authorized/approved Covid-19 primary series vaccine with or without an mRNA-based booster as the third dose in the series ≥90 days prior to screening. Participants who had previously received 2 injections of any Covid-19 vaccine, including a mixed regimen, were eligible to receive mRNA-1273.529, mRNA-1273.214, or mRNA-1273 as the first booster (third) dose, and those who previously received an mRNA Covid-19 first booster (third) dose were eligible to receive these vaccines as the second booster (fourth) dose. Participants who had a history of positive SARS-CoV-2 testing within 90 days of screening were ineligible for the study (inclusion/exclusion criteria and study design are detailed in the Supplementary Appendix).

The study was conducted in accordance with the International Council for Harmonisation of Technical Requirements for Registration of Pharmaceuticals for Human Use, Good Clinical Practice guidelines. The Derby Research Ethics Committee approved the protocol and consent forms. All participants provided written informed consent. The sponsor was involved in the study design as well as the collection, analysis, and interpretation of the data. The authors vouch for the completeness and accuracy of the data, for the fidelity of the study to the protocol, and made the decision to submit the manuscript for publication.

### Trial procedures

The monovalent mRNA-1273 and mRNA-1273.529 vaccines contain a single mRNA (50 µg) encoding the prefusion stabilized S glycoprotein of the ancestral (Wuhan-Hu-1; mRNA-1273) or the omicron BA.1 variant (mRNA-1273.529), respectively. Bivalent mRNA-1273.214 contains 2 mRNAs at a 1:1 ratio (25-μg each for a total of 50 µg) encoding the prefusion-stabilized S glycoprotein of ancestral SARS-CoV-2 (Wuhan-Hu-1) and the omicron BA.1 variant. Vaccines (50 µg) were administered in a 0.25-0.5 ml volume as an intramuscular injection into the deltoid muscle on day 1.

Participants were randomized in a 1:1 ratio to receive a single dose of either mRNA-1273.529 (50 µg) or mRNA-1273 (50 µg) in part 1 of the study and to receive either mRNA-1273.214 (50 µg) or mRNA-1273 (50 µg) in part 2 of the study. In both study parts, randomization was stratified by age group (16-<65 or ≥65 years) and the number of prior booster doses received (zero or one).

### Objectives

The primary objective of both study parts was to evaluate the safety and reactogenicity of booster doses of mRNA-1273.529 (part 1), mRNA-1273.214 (part 2), and mRNA-1273 (parts 1 and 2). Immunogenicity objectives of part 1 were to demonstrate non-inferiority (primary) or superiority (key secondary) of mRNA-1273.529-elicited immune responses to omicron BA.1 at day 29 compared with mRNA-1273 when administered as a booster dose. Additional part 1 secondary objectives include noninferiority of the immune response of mRNA-1273.529 compared to mRNA-1273 booster against ancestral SARS-CoV-2 with the D614G mutation (ancestral SARS-CoV-2 [D614G]) at the day 29 and the seroresponse (SRR) of mRNA-1273.529 and mRNA-1273 boosters administered as booster doses. Incidence of symptomatic and asymptomatic SARS-CoV-2 infection was an exploratory objective.

Primary immunogenicity objectives of part 2 were to demonstrate the non-inferiority of mRNA-1273.214-elicited immune responses to omicron BA.1 and to the ancestral SARS-CoV-2 (D614G), as well as superiority of mRNA-1273.214-elicited immune responses to omicron BA.1, at day 29 compared with mRNA-1273 when administered as booster doses. Part 2 secondary objectives included evaluating the immune response of mRNA-1273.214 against other variant strains at day 29, the SRR of mRNA-1273.214 and mRNA-1273, and symptomatic and asymptomatic SARS-CoV-2 infection after mRNA-1273.214 or mRNA-1273 booster vaccination. Part 1 and 2 objectives are detailed in the Supplementary Appendix and Table S1.

### Safety Assessments

Safety assessments included solicited local and systemic adverse reactions (ARs) recorded within 7 days after booster vaccination; unsolicited adverse events (AEs) within 28 days after vaccination; and serious AEs (SAEs), medically attended AEs (MAAEs), AEs leading to withdrawal, and AEs of special interest (AESIs) from vaccination to end of study.

### Immunogenicity Assessments

Immunogenicity was assessed by geometric mean concentrations (GMCs) of serum neutralizing antibodies against omicron BA.1 and/or SARS-CoV-2 (ancestral SARS-CoV-2 (D614G]) as measured by validated pseudovirus neutralization assays (PsVNA).^15,16^ GMCs of serum binding antibodies against SARS-CoV-2 (ancestral or omicron BA.1) were determined using a validated binding assay (Meso Scale Discovery) to the SARS-CoV-2-specific S protein in part 2 of the study. The seroresponse (SRR) against SARS-CoV-2 ancestral (D614G) and omicron BA.1 after vaccination were also evaluated. Immunogenicity assessment and assays are further detailed in the Supplementary Appendix.

### Incidence of SARS-CoV-2 Infections

The number and incidence rates per 1000 person years of Covid-19 disease per the primary definition in the coronavirus efficacy (COVE) trial and per the CDC definition,^17-19^ SARS-CoV-2 infection regardless of symptoms, as well as asymptomatic SARS-CoV-2 infection were assessed for each study arm (supplementary methods). Active surveillance for Covid-19 and SARS-CoV-2 infection were performed in both parts of the study (Supplementary methods). SARS-CoV-2-infection status is defined on the basis of either bAb levels against SARS-CoV-2 nucleocapsid (Roche Elecsys) or RT-PCR test that become positive post-baseline. In this interim analysis, viral variant sequences were obtained by RT-PCR from nasopharyngeal swabs of participants positive for SARS-CoV-2-infection from 14 days after the study vaccine administration cumulatively on an ongoing basis. Sequences identified in Covid-19 cases were assessed through the data cutoff date.

### Statistical Analyses

Statistical analysis methods are detailed in the supplementary appendix and analysis sets in Table S2. Safety was assessed in the safety set (all participants who received vaccination) with analyses of solicited ARs performed in the solicited safety set (participants in the safety set who contributed solicited AR data). The per-protocol set for immunogenicity (PPSI) consists of participants in the full analysis set who received the planned booster dose, had pre-booster and day 29 antibody data available and no major protocol deviations. The primary immunogenicity objectives were assessed in the PPSI-SARS-CoV-2 negative set (PPSI-negative) in this interim analysis (Fig. S2). The incidences of SARS-CoV-2 infection were evaluated in the per-protocol set for efficacy (PPSE) comprising all participants in the modified-intent-to-treat population who received the planned study vaccination and had no major protocol deviations. The immunogenicity and efficacy objectives were evaluated only in those who received second booster (4^th^) doses; those who received 1^st^ booster (3^rd^) doses in the study were excluded from these analyses.

For immunogenicity analyses, the GMC of neutralizing and binding antibodies was calculated at day 29 with corresponding 95% confidence intervals (CIs) for each vaccine group. The geometric mean fold-rise (GMFR) of post-booster/pre-booster concentrations with 95% CIs at day 29 is provided. The SRRs summarized at day 29 for each treatment group with the 95% CI (Clopper-Pearson) are provided. The difference of SRRs at day 29 for mRNA-1273.529 and mRNA-1273.214 compared with mRNA-1273 with 95% CI (Miettinen-Nurminen method) are provided. In part 1, the primary immunogenicity objective, non-inferiority of the antibody responses against the omicron BA.1 following mRNA-1273.529 compared to mRNA-1273 boosters at day 29 is considered met if the lower bound of the 99% CI of the GMC ratio (GMR) of mRNA-1273.529 vs. mRNA-1273 is ≥0.67 based on a non-inferiority margin of 1.5 at a 2-sided alpha of 0.01. Dependent on non-inferiority being demonstrated, superiority of mRNA-1273.529 versus mRNA-1273 against omicron BA.1 was subsequently tested and considered demonstrated if the lower bound of the 99% CI of the GMR was >1 at day 29. Non-inferiority of the antibody responses against ancestral SARS-CoV-2 (D614G) following mRNA-1273.529 compared to mRNA-1273 booster at day 29 is considered met if the lower bound of the 95% CI for GMR is ≥0.67. The primary immunogenicity objectives in part 2 were evaluated by hypothesis testing using a prespecified testing sequence (Fig. S3 and detailed in the Supplementary Appendix). Non-inferiority of mRNA-1273.214 versus mRNA-1273 against both the omicron BA.1 and the ancestral SARS-CoV-2 (D614G) at day 29 was assessed using a non-inferiority margin of 1.5 at 2-sided alpha of 0.01 and was considered demonstrated if the lower bound of the 99% CI of the GMR (mRNA-1273.214 vs mRNA-1273) against omicron BA.1 and ancestral SARS-CoV-2 (D614G) was ≥0.67. Superiority of mRNA-1273.214 versus mRNA-1273 against omicron BA.1 at day 29 was evaluated if non-inferiority objectives were met and was demonstrated if the lower bound of the GMR ruled out 1 (>1) at day 29.

The number and percentage of participants with SARS-CoV-2 infection and Covid-19 events starting at 14 days after randomization are summarized. Cumulative event rates were calculated using a Kaplan-Meier method where time to event is calculated as the time starting 14 days after randomization. The incidence rates of Covid-19 cases adjusting for person-time and 95% CIs used an exact method (Poisson distribution) were summarized. The relative vaccine efficacy (VE) is estimated by a proportional hazards model in the mRNA-1273.214 group compared to mRNA-1273. In an exploratory analysis, Covid-19 cases having variant sequences (BA.2, BA.4, and BA.5 lineages) were assessed using a competing risk method to analyze sublineage-specific events, where competing events were not censored. The Fine-Gray proportional hazards model for the subdistribution of a competing risk was used to estimate the hazard ratio and relative vaccine efficacy VE (1-hazard ratio).^20^

All analyses were conducted using SAS Version 9.4 or higher.

## Results

### Study population

Between February 16, 2022 and March 24, 2022, 724 eligible participants were randomized in part 1 of the study and between April 2, 2022 and June 17, 2022, 2,824 participants were randomized in part 2 of the study (Fig. 1). Of these, 719 participants in part 1 received mRNA-1273.529 (n=362) or mRNA-1273 (n=357) and in part 2, 2824 received mRNA-1273.214 (n=1418) or mRNA-1273 (n=1395) in the full analysis set.

**Fig. 1.**
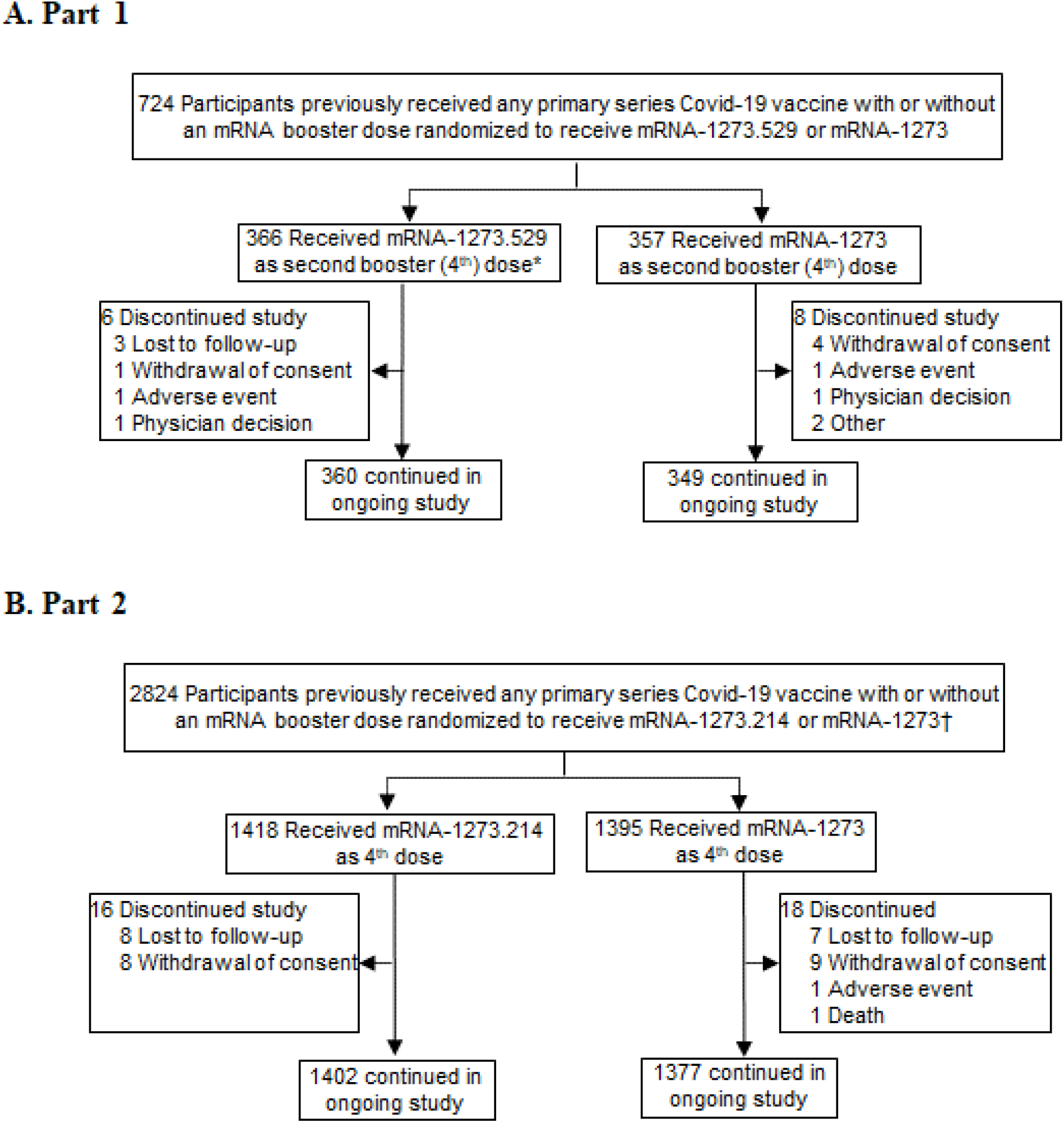
Trial Profile. Eligible participants who previously received any Covid-19 vaccine primary series with or without a prior booster dose, were randomized in a 1:1 ratio to receive either a mRNA-1273.529 or a mRNA-1273 booster in part 1 or were randomized in a 1:1 ratio to receive either a mRNA-1273.214 or a mRNA-1273 booster in part 2. Randomization was stratified by age group (16 to <65 or ≥65 years) and number of prior booster doses received (received study vaccine as the 4th dose or as the 3rd dose). Participants who received the second booster (4^th^) dose as part of the study must have previously received a mRNA vaccine as the first booster (3^rd^) dose of a Covid-19 vaccine. Participants who received the first booster (3^rd^) dose may have previously received 2 injections of an approved/authorized mRNA or non-mRNA Covid-19 vaccine. *In part 1, 4 participants who received mRNA-1273.529 were included in the safety set but not in the full analysis set, and 1 participant received mRNA-1273.529 as a 3^rd^ dose and was excluded from the immunogenicity and efficacy analyses. †In part 2, 4 participants received mRNA-1273.214 as a 3rd dose and 7 participants received mRNA-1273 as a 3rd dose and were excluded from the immunogenicity and efficacy analyses. The data-cutoff date was August 4, 2022.

Baseline characteristics between study arms in both study parts were generally balanced (Table 1). The mean age of participants was 57 years (range 19-87 in part 1; 17-89 in part 2) in all study arms of the study. In both study parts, ∼33% of the participants were ≥65 year of age, ∼50% were female, and the majority were white (∼93%). The proportions of participants with prior SARS-CoV-2-infection pre-booster were 12.8% and 12.0% in the mRNA-1273.529 and mRNA-1273 arms, and 22.6% and 26.0% in the mRNA-1273.214 and mRNA-1273 arms, respectively. Median duration times (months [interquartile ranges]) between the most recent Covid-19 vaccine and study boosters were similar in the mRNA-1273.529 (4.0 [3.6-4.7]) and mRNA-1273 (4.1 [3.5-4.7]) arms and in the mRNA-1273.214 (5.5 [4.8-6.2] and mRNA-1273 (5.4 [4.8-6.2]) arms. The majority of participants received the Covid-19 vaccines Vaxzevria (52% and 63%) or Comirnaty (46% and 34%) as primary vaccination series, and Comirnaty (81% and 77%) as the first booster vaccine in parts 1 and 2, respectively (Table 1 and Fig. S4).

**Table 1:** Demographics and Participant Characteristics, Safety Set.

| Characteristics n (%)<sup>*</sup> | Part 1 |  | Part 2 |  |
| --- | --- | --- | --- | --- |
|  | mRNA-1273.529<br>50 µg<br>N=367 | mRNA-1273<br>50 µg<br>N=357 | mRNA-1273.214<br>50 µg<br>N=1422 | mRNA-1273<br>50 µg<br>N=1402 |
| Age at Screening (yr)<br>Mean (range) | 57.6 (19-87) | 57.3 (21-87) | 57.4 (18-89) | 57.0 (17-81) |
| Age subgroup |  |  |  |  |
| ≥16 and <65 years | 240 (65.4) | 233 (65.3) | 945 (66.5) | 933 (66.5) |
| ≥65 years | 127 (34.6) | 124 (34.7) | 477 (33.5) | 469 (33.5) |
| Sex |  |  |  |  |
| Male | 167 (45.5) | 155 (43.4) | 727 (51.1) | 708 (50.5) |
| Female | 200 (54.5) | 202 (56.6) | 695 (48.9) | 694 (49.5) |
| Race or ethnic group<sup>†</sup> |  |  |  |  |
| White | 353 (96.2) | 335 (93.8) | 1346 (94.7) | 1312 (93.6) |
| Black | 0 | 0 | 6 (0.4) | 6 (0.4) |
| Asian | 10 (2.7) | 10 (2.8) | 31 (2.2) | 41 (2.9) |
| Mixed or multiple ethnic groups | 3 (0.8) | 5 (1.4) | 21 (1.5) | 28 (2.0) |
| Other | 1 (0.3) | 5 (1.4) | 4 (0.3) | 7 (0.5) |
| Not reported, unknown or missing | 0 | 2 (0.6) | 14 (1.0) | 8 (0.6) |
| Body mass index (kg/m<sup>2</sup>)<br>Mean (SD) | 27.7 (5.7) | 28.3 (5.8) | 27.7 (5.5) | 27.6 (5.7) |
| Time from most recent Covid-19 vaccine to<br>booster dose (months)<br>Median (interquartile range)<sup>§</sup> | 4.0 (3.6-4.7) | 4.1 (3.5-4.7) | 5.5 (4.8-6.2) | 5.4 (4.8-6.2) |
| Prior vaccination received (primary series) |  |  |  |  |
| Vaxzevria | 196 (53.4) | 178 (49.9) | 907 (63.8) | 872 (62.2) |
| Comirnaty | 164 (44.7) | 170 (47.6) | 472 (33.2) | 493 (35.2) |
| Spikevax | 2 (0.5) | 2 (0.6) | 21 (1.5) | 19 (1.4) |
| Jcovden | - | - | 4 (0.3) | 5 (0.4) |
| Mixed regimen/other | 5 (1.3) | 7 (2.0) | 18 (1.3) | 13 (0.9) |
| Prior first booster received |  |  |  |  |
| Comirnaty | 296 (80.9) | 290 (81.2) | 1110 (78.1) | 1068 (76.6) |
| Spikevax | 70 (19.1) | 67 (18.8) | 308 (21.7) | 327 (23.5) |
| Pre-booster RT-PCR assay for SARS-CoV-2 |  |  |  |  |
| Negative | 363 (98.9) | 351 (98.3) | 1302 (91.6) | 1291 (92.1) |
| Positive | 4 (1.1) | 6 (1.7) | 19 (1.3) | 15 (1.1) |
| Missing | 0 | 0 | 101 (7.1) | 96 (6.8) |
| Pre-booster antibody to SARS-CoV-2<br>nucleocapsid<sup>¶</sup> |  |  |  |  |
| Negative | 319 (86.9) | 318 (89.1) | 1093 (76.9) | 1034 (73.8) |
| Positive | 44 (12.0) | 37 (10.4) | 313 (22.0) | 357 (25.5) |
| Missing | 4 (1.1) | 2 (0.6) | 16 (1.1) | 11 (0.8) |
| Pre-booster SARS-CoV-2 status |  |  |  |  |
| Negative | 316 (86.1) | 312 (87.4) | 1004 (70.6) | 957 (68.3) |
| Positive | 47 (12.8) | 43 (12.0) | 322 (22.6) | 364 (26.0) |
| Missing | 4 (1.1) | 2 (0.6) | 96 (6.8) | 81 (5.8) |

Percentages are based on the number of participants in the safety set.
<sup>\*</sup>Percentages may not total 100% due to rounding. RT-PCR: reverse-transcriptase polymerase chain reaction; SARS-CoV-2: severe acute respiratory syndrome coronavirus 2.
<sup>†</sup>Race and ethnic group were reported by the participant.
<sup>§</sup>Participants with <3 months duration between 3rd and 4th doses were excluded from the per-protocol sets
<sup>¶</sup>The Elecsys assay for binding antibody to SARS-CoV-2 nucleocapsid was used.
<sup>||</sup>Pre-booster SARS-CoV-2 status was positive if there was evidence of previous SARS-CoV-2 infection, defined as positive binding antibody against the SARS-CoV-2 nucleocapsid or positive RT-PCR assay at day 1; negative SARS-CoV-2 status was defined as negative binding antibody against the SARS-CoV-2 nucleocapsid and a negative RT-PCR assay at day 1. The data-cutoff date was August 4, 2022.

### Immunogenicity

In the part 1 primary immunogenicity analysis in participants in the PPSI-negative set who had no evidence of SARS-CoV-2 infection up to the day of the analysis visit, the observed neutralizing antibody GMCs (95% CI) against omicron BA.1 were higher following the mRNA-1273.529 (537.7 [478.2−604.6]) booster at 28 days than after the mRNA-1273 (307.4 [279.5−338.2]) booster (Table 2, Fig. S5 and Supplementary Appendix). Neutralizing antibody GMCs (99% CI) estimated by an ANCOVA model against omicron BA.1 were 525.5 (472.0−585.0) and 312.8 (281.4−347.7) 28 days after the mRNA-1273.529 and mRNA-1273 boosters, respectively, with a GMR (99% CI) of 1.68 (1.45−1.95), which met the pre-specified criterion of non-inferiority (lower bound of CI ≥0.67) (Table 2). Additionally, superiority of the immune response of mRNA-1273.529 compared to the mRNA-1273 booster against omicron BA.1 (key secondary objective) was demonstrated (lower bound of the CI >1). The omicron-BA.1 SRRs (95% CI) were 82.7% (77.6−87.1%) and 55.7% (49.6−61.7%) 28 days after the mRNA-1273.529 and mRNA-1273 booster doses, respectively, and the SRR (95% CI) difference was 27.0 (19.4−34.3).

**Table 2.**
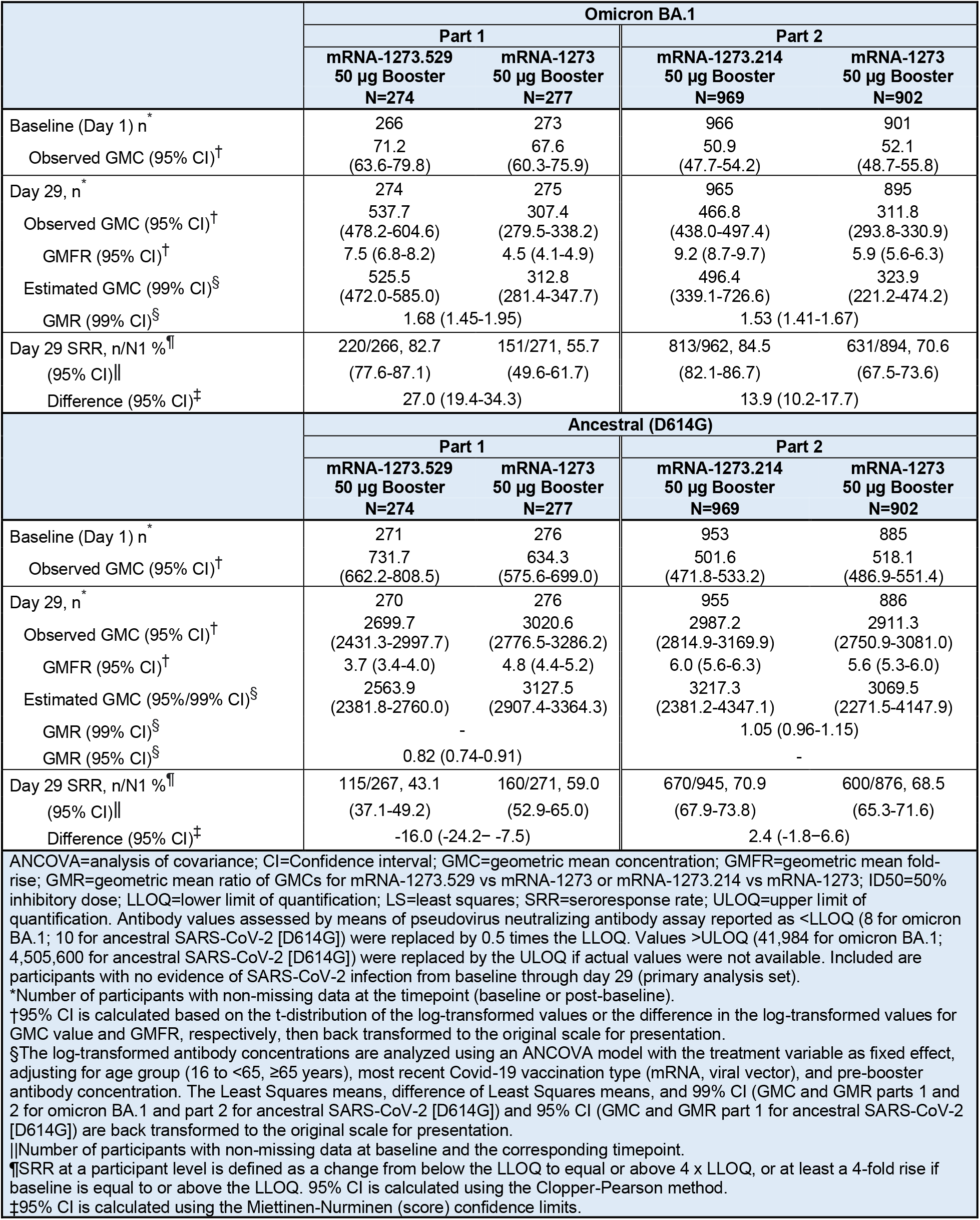
Pseudovirus Neutralizing Antibodies Against Omicron BA.1 or Ancestral SARS-CoV-2 (D614G) after Receipt of 50-µg of mRNA-1273.529, mRNA-1273.214, or mRNA-1273 Boosters Administered to Participants with No Prior SARS-CoV-2-Infection.

In the secondary analysis of immunogenicity in the part 1 PPSI-negative set, the observed ancestral SARS-CoV-2 (D614G) GMCs (95% CI) were 2699.7 (2431.3−2997.7) for the mRNA-1273.529 and 3020.6 (2776.5−3286.2) for the mRNA-1273 boosters at day 29. The estimated neutralizing antibody GMCs (95% CI) were 2563.9 (2381.8−2760.0) and 3127.5 (2907.4−3364.3) in the mRNA-1273.529 and mRNA-1273 arms, respectively, with a GMR (95% CI) of 0.82 (0.74−0.91) at day 29, meeting the secondary non-inferiority objective (95% CI lower bound ≥0.67). The ancestral SARS-CoV-2 (614G) SRRs (95% CI) were 43.1% (37.1−49.2%) and 59.0% (52.9−65.0%) in the mRNA-1273.529 and mRNA-1273 arms, respectively (SRR difference -16.0 [95% CI, -24.2−-7.5]).

In part 2, in the primary analysis in the PPSI-negative set, the observed neutralizing antibody GMCs (95% CIs) against omicron BA.1 were also higher (466.8 [438.0−497.4]) after the mRNA-1273.214 than the mRNA-1273 (311.8 [293.8−330.9]) booster, at day 29. The estimated GMCs (99% CI) against omicron BA.1 were 496.4 (339.1-726.6) and 323.9 (221.2-474.2), 28 days following the mRNA-1273.214 and mRNA-1273 boosters, respectively with a GMR (99% CI) of 1.53 (1.41−1.67), which met the pre-specified primary immunogenicity criteria of non-inferiority (lower bound of CI ≥0.67) and superiority (lower bound of CI >1) (Table 2). For the part 2 co-primary endpoint, the observed ancestral SARS-CoV-2 (D614G) GMCs were 2987.2 (2814.9−3169.9) after the mRNA-1273.214 and 2911.3 (2750.9−3081.0) after the mRNA-1273 boosters at day 29. The estimated GMCs (99% CI) were 3217.3 (2381.2−4347.1) and 3069.5 (2271.5−4147.9), 28 days after the mRNA-1273.214 and mRNA-1273 booster doses, respectively with a GMR (99% CI) of 1.05 (0.96-1.15), which met the pre-specified criterion of non-inferiority (lower bound of CI ≥0.67). The SRRs (95% CI) were 84.5% (82.1−86.7%) and 70.6% (67.5−73.6%) for omicron BA.1 (SRR difference 13.9 [10.2−17.7]), and 70.9% (67.9−73.8%) and 68.5% (65.3−71.6%) for ancestral SARS-CoV-2 (D614G) (SRR difference of 2.4 [-1.8−6.6], 28 days after the mRNA-1273.214 and mRNA-1273 booster doses, respectively. In the immunogenicity cohorts containing all participants regardless of SARS-CoV-2-infection status and only those with evidence of prior SARS-CoV-2-infection, GMCs were also higher following the mRNA-1273.214 than mRNA-1273 booster against both omicron BA.1 and ancestral SARS-CoV-2 (D614G) (Fig. S6 and Table S3).

In part 1, in the PPSI-negative set, spike binding antibody GMCs were higher against omicron BA.1 after the mRNA-1273.529 than mRNA-1273 booster at day 29 (GMR 1.18 [95% CI, 1.05−1.32]) and were similar against ancestral SARS-CoV-2 (D614G) and alpha, gamma, and delta variants with GMRs (95% CI) ranging from 0.91 (0.84−0.99) to 1.00 (0.92−1.08) for both boosters (Table S4 and Fig. S7). In part 2, estimated GMCs were higher after the mRNA-1273.214 than after the mRNA-1273 booster across omicron BA.1, ancestral SARS-CoV-2 (D614G) and alpha, gamma, and delta variants (Table S5 and Fig. S8). The GMRs (95% CI) ranged from 1.06 (1.01−1.11) to 1.13 (1.06−1.21) across the variants.

### Safety

The median (range) safety follow-up times were 156 (29-170) days for both mRNA-1273.529 and mRNA-1273 in part 1, and 102 (6-125) days for both mRNA-1273.214 and mRNA-1273 in part 2. The percentage of participants reporting solicited local and systemic adverse reactions within 7 days after the booster dose was similar in the part 1 mRNA-1273.529 (91.3%) and mRNA-1273 (93.3%) groups and in the part 2 mRNA-1273.214 (90.4%) and mRNA-1273 (94.2%) (Fig. 2 and Table S6). The most commonly reported adverse reactions in both parts 1 and 2 were pain, fatigue, and headache. The majority of reactions were grades 1-2. Incidences of grade 3 events were similar across study arms, and no grade 4 events were reported.

**Fig. 2.**
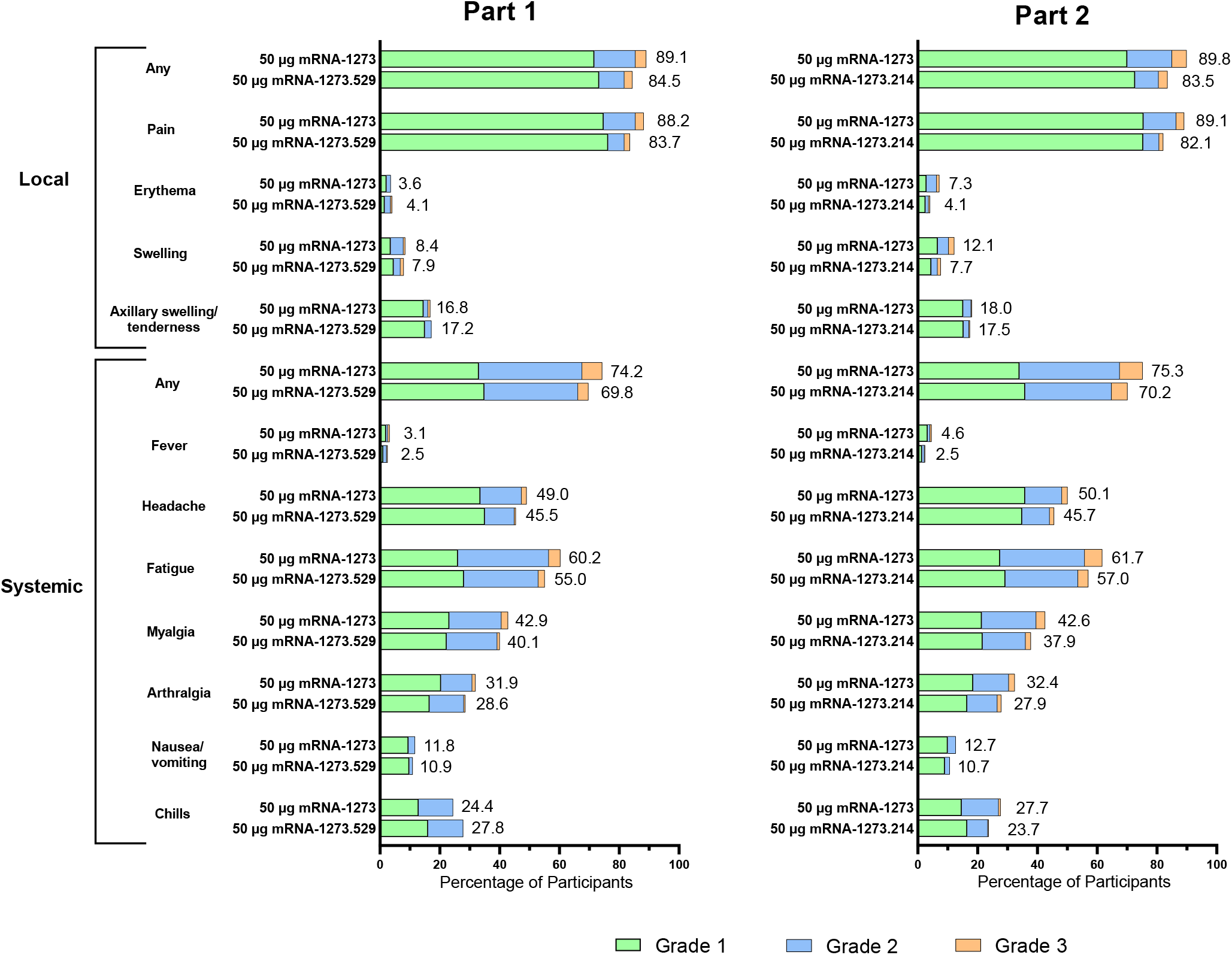
Solicited Adverse Reactions after Receipt of mRNA-1273.214, mRNA-1273.529, or mRNA-1273 Boosters, Safety Set. Shown are the percentages of participants in whom solicited local or systemic adverse reactions occurred within 7 days after the booster dose in the solicited safety set (part 1, n=367 in the mRNA-1273.529 and n=357 in the mRNA-1273 groups; part 2, n=1421 in the mRNA-1273.214 and 1398 in the mRNA-1273 groups).

In part 1 of the study, the percentages of participants reporting any unsolicited AEs within 28 days after the booster dose were similar in the mRNA-1273.529 (39.2%) and mRNA-1273 (35.6%) groups (Table S7). The overall incidences of AEs considered related to study vaccine by the investigators were 27 (7.4%) in the mRNA-1273.529 and 30 (8.4%) in the mRNA-1273 groups. Rates of related medically-attended AEs were low in the mRNA-1273.529 (5 [1.4%]) and mRNA-1273 groups (2 [0.6%]). In the mRNA-1273.529 arm, 5 SAEs occurred in 5 participants, and 1 SAE occurred in 1 participant in the mRNA-1273 arm. Of these, 2 SAEs (bilateral pulmonary emboli [PE] and ventricular tachycardia [VT]) in the mRNA-1273.529 arm were considered related to study vaccine. The SAE of PE led the participant to discontinue from the study; this was the only AE that led to study discontinuation in part 1.

In part 2, the frequencies of any unsolicited adverse events reported within 28 days after the booster dose were also similar in the mRNA-1273.214 (31.5%) and mRNA-1273 (29.7%) groups (Table S7). The incidences of AEs considered related to study vaccine by the investigators were 69 (4.9%) and 71 (5.1%) in the mRNA-1273.214 and mRNA-1273 groups respectively. Related medically-attended AEs occurred in 6 (0.4%) and 7 (0.5%) in the mRNA-1273.214 and mRNA-1273 groups, respectively. None of the AEs led to study discontinuation. Serious AEs occurred in 6 (0.4%) participants in the mRNA-1273.214 group and 5 (0.4%) in the mRNA-1273 group; none were considered by the investigator to be related to the study vaccine among mRNA-1273.214 recipients. One SAE (multiple pulmonary emboli) was considered related to the study vaccine by the investigator in the mRNA-1273 control arm.

As of the interim analysis data cutoff date, there were no fatal AEs. After the interim analysis data cut-off date, two deaths (sudden unexpected death in epilepsy, sudden cardiac death due to arrhythmia) occurred in participants in the mRNA-1273 arm of part 2 and were determined by the investigators to be unrelated to the study vaccine.

### Incidence of SAR-CoV-2 Infection

The total overall person years at time of the data cut-off were 113.3 and 111.8 in the mRNA-1273.529 and mRNA-1273 arms, respectively, in part 1 of the study (Table S8). In an analysis of the part 1 exploratory objective, the incidence rates (95% CI) of Covid-19 based on the primary case definition used in the COVE trial^18,19^ 14 days after randomization were 670.5 (528.3−839.3) and 769.3 (615.4−950.1) per 1000 person-years, in the mRNA-1273.529 and mRNA-1273 arms, respectively (Table S8 and Fig. 3). Incidence rates (95% CI) starting 14 days after randomization for Covid-19 based on the CDC definition^17^ were 731.1 (580.6−908.7) and 888.3 (720.4−1083.7) and for overall SARS-CoV-2 infection were 862.5 (696.2−1056.7) in the mRNA-1273.529 and 1012.9 (830.1−1224.0) in mRNA-1273 groups per 1000 person-years, respectively. One Covid-19-related hospitalization occurred in part 1 in a participant who received mRNA-1273.529, diagnosed using a rapid antigen/lateral flow test and not using a PCR-based test.

**Fig. 3.**
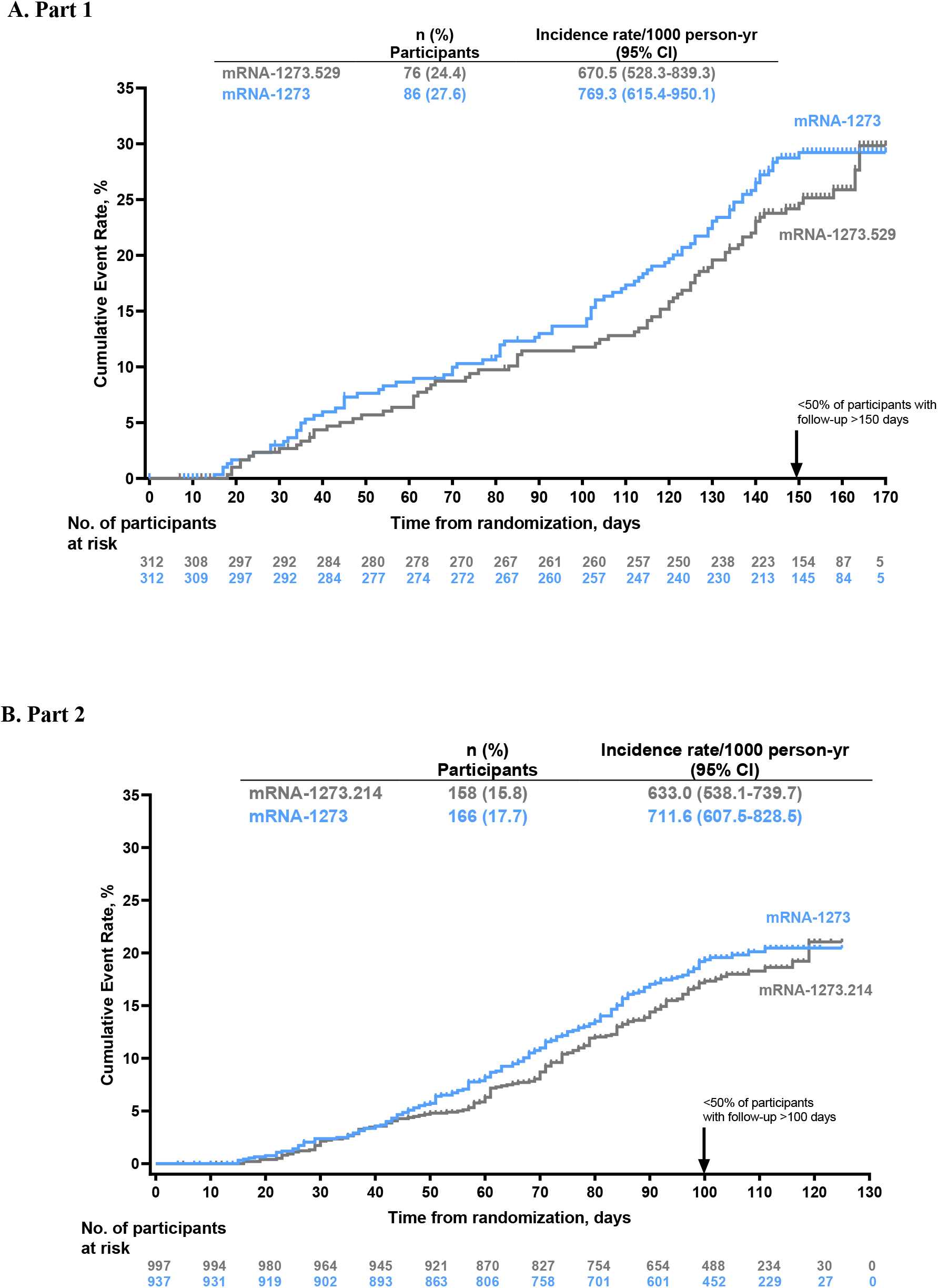
Cumulative Event Rates of Covid-19 Starting ≥14 Days After Randomization Following Receipt of mRNA-1273.529, mRNA-1273.214, or mRNA-1273 Boosters. Shown are the cumulative event rates of Covid-19 based on assessment starting 14 days after randomization in the per-protocol efficacy population of parts 1 (Panel A) and 2 (Panel B). Tick marks indicate censored data. The incidence rate was defined as the number of events divided by number of participants at risk and was adjusted by person-years. Arrow denotes that as of the data cutoff date for the interim analysis <50% of participants had follow-up beyond 150 days in part 1 and beyond 100 days in part 2.

In part 2, the total overall person years at time of the data cut-off were 240.5 and 225.6 in the mRNA-1273.214 and mRNA-1273 arms, respectively (Table S8). In an analysis of the secondary part 2 objective, the estimated incidence rates (95% CI) of Covid-19 based on the COVE primary case definition^18,19^ starting 14 days after randomization were 633.0 (538.1−739.7) in the mRNA-1273.214 and 711.6 (607.5−828.5) in the mRNA-1273 arms per 1,000 person years (Table S8 and Fig. 3). The incidence rates (95% CI) starting 14 days after randomization were 739.2 (635.7−854.8) and 755.4 (647.6−876.0) for Covid-19 based on the CDC definition^17^ and for overall SARS-CoV-2 infection were 1010.5 (887.5−1145.9) and 1099.1 (966.5−1244.7) in the mRNA-1273.214 and mRNA-1273 arms, respectively. No cases of Covid-19-related hospitalization were reported in part 2.

Due to the evolving omicron subvariant landscape, RNA sequence information was obtained by RT-PCR from nasopharyngeal swabs positive for SARS-CoV-2-infection collected from February 2022 through September 2022. Of the total 850 available variant sequences in parts 1 and 2, the majority were of the omicron BA.4 and BA.5 lineages and were predominant during June and July of 2022 (Fig. S9). Of the total 324 Covid-19 cases in part 2 that occurred ≥14 post-randomization during this interim analysis period, variant sequences were detected among 135 and 154 of the cases in the mRNA-1273.214 and mRNA-1273 arms, respectively; the majority (70% and 60%) were of the omicron BA.5, 13% and 19% were of the BA.4, and 17% and 21% were of the BA.2 lineages (Table S9).

An exploratory analysis of Covid-19 cases having sequences of the BA.2, BA.4, and BA.5 sublineages using the Fine-Gray proportional hazards model for the subdistribution of a competing risk,^20^ showed a lower incidence rate in the mRNA-1273.214 arm compared with mRNA-1273 for the BA.2 and BA.4 sublineages; this trend was not seen for the BA.5 lineage group (Fig. S10 and Table S10). The relative VE estimates (95% CI) for mRNA-1273.214 vs. mRNA-1273 were 32.6% (−15.1−60.5%), 41.6% (−5.1−67.5%), and 4.4% (−27.2−28.2%) for the BA.2, BA.4, and BA.5 sublineages, respectively. A sensitivity analysis of the relative vaccine efficacy against variant sublineages excluding BA.5 sublineages resulted in a relative VE of 37.3% and a 95% CI excluding zero (6.9−57.8%).

## Discussion

Previous studies have indicated that bivalent booster vaccines containing omicron variants elicit higher neutralizing antibody responses compared to original boosters.^1-3^ The present study is the first large randomized, observer-blind, active-controlled, phase 3 clinical trial comparing head-to-head variant-targeting mRNA-1273 booster vaccines with the original mRNA-1273 vaccine. The results demonstrate that the bivalent mRNA-1273.214 (25-μg omicron BA.1 and 25-μg ancestral SARS-CoV-2) vaccine elicited superior neutralizing antibody responses against omicron BA.1 and non-inferior responses against ancestral SARS-CoV-2 (D614G), compared to 50 μg of mRNA-1273 28 days after the booster dose, which is consistent with previously published results.^1-3,21^ Additionally, we compared mRNA-1273.529 50 μg, a monovalent omicron BA.1 booster, with mRNA-1273 50 μg, and mRNA-1273.529 also elicited superior neutralizing antibody responses against omicron BA.1 compared to mRNA-1273. However, the ancestral SARS-CoV-2 (D614G) neutralizing and binding antibody concentrations and SRRs as well as the alpha, gamma, and delta variant binding antibody responses for the monovalent omicron BA.1 booster mRNA-1273.529 were similar compared to mRNA-1273, suggesting a more restricted immune response with monovalent variant-targeting booster vaccines.^22,23^

The distinct study design as well as the disparate laboratory vendor and immunoassays utilized may explain differences in the absolute value of the geometric mean antibody concentrations observed in this study compared to our open-label study of mRNA-1273.214.^1^ In the present study, most participants received a non-mRNA-based Covid-19 primary series vaccination, and the primary immunogenicity analysis included only those who had no evidence of SARS-CoV-2 infection from baseline until the day 29 analysis. This contrasts with our open-label, non-randomized study where participants had only previously received mRNA-1273 and may have had SARS-CoV-2 infection between study vaccination and day 29. Also of note, the lower SRR in this study is likely due to measurement of changes in antibody levels at day 29 from pre-booster levels rather than changes from pre-primary series levels used in other studies.^1-3^ Despite these differences in study design, consistent results of superior antibody responses against omicron BA.1 and non-inferior responses against the ancestral SARS-CoV-2 were observed with the bivalent mRNA-1273.214 booster.

Emerging observational data suggest a clinical benefit in preventing Covid-19 with the use of bivalent booster vaccines in the setting of emergent variants beyond omicron BA.1.^7-9,24^ Although this study was not sufficiently powered to detect a difference in relative vaccine efficacy compared to mRNA-1273, a numerically lower incidence rate of Covid-19 was observed with mRNA-1273.214 versus mRNA-1273. The trend was notable when considering Covid-19 events caused by omicron BA.2 and BA.4 isolates but was not the case for BA.5 isolates. The spike protein sequences between omicron BA.4 and BA.5 are identical and neutralization results are typically reported in the same assay and reported together for both sublineages (BA.4/BA.5).^1-3,11,12^ Factors that may explain the different relative VE point estimates between the omicron BA.4 and BA.5 sublineages include the increased viral fitness of BA.5 compared to other sublineages (including BA.4), the later timing of the BA.5 emergence in relation to vaccination in the study when antibody titers may have begun to wane, and the fact that the study was underpowered to detect differences in relative VE in the limited amount of follow-up time for this interim analysis. Overall, the results of the post-booster Covid-19 incidence rates suggest a clinical benefit of the bivalent vaccine despite the viral evolution to variant sublineages not contained in the mRNA-1273.214 vaccine. SARS-CoV-2 continues to evolve and omicron sublineages such as BQ.1.1, XBB.1, and XBB.1.5 have emerged with spike mutations that can confer antibody escape leading to re-infections and Covid-19.^11,12,25^ In this setting of emerging divergent variants, it is important to continue monitoring vaccine effectiveness in real-world studies in parallel with assessing the cross-neutralization ability of previously authorized vaccines.

The incidences of solicited adverse reactions with mRNA-1273.529 and mRNA-1273.214 were similar to that of mRNA-1273, which is consistent with previously published results.^1-3^ No new safety concerns were identified in this interim analysis up to the day 29 data cutoff date. The safety data presented here, taken together with the previously reported longer-term safety follow-up on the BA.1 bivalent booster mRNA-1273.214^3^ extend the body of safety information available for omicron-targeting bivalent vaccines. Longer-term evaluation of the safety of mRNA-1273.214 and mRNA-1273.529 continues in this ongoing study.

Real-world vaccine effectiveness data previously indicated a decrease in the original vaccine effectiveness in the setting of divergent SARS-CoV-2 variants and omicron sublineages in particular.^7-9^ The potential added benefit of variant-containing mRNA-1273 booster vaccines versus the original mRNA-1273 vaccines in protecting against Covid-19 had not been demonstrated in a large, randomized comparative trials. Additionally, the rapidly shifting epidemiology of SARS-CoV-2 precluded large trials that would evaluate a vaccine containing the circulating variant while the same variant was still epidemiologically dominant. Therefore, regulatory decision-making for vaccine updates was based on preclinical information and smaller, open-label, non-randomized clinical studies with results from such studies becoming available post-authorization. The present study was designed to address the need to rapidly respond to the emergence of the omicron BA.1 variant as a randomized trial, initiated in February 2022 with interim results (to August 4, 2022 data cutoff date) becoming available in late November 2022. We evaluated omicron BA.1-containing vaccines versus the original mRNA-1273 vaccine, and when the regulatory decision was made to update the vaccine with omicron BA.4/BA.5, it was no longer feasible to randomize additional participants to evaluate the newer bivalent vaccine. The study was powered to detect differences in immunogenicity between the two vaccines but not in Covid-19 rates post-boost and Covid-19 events are subject to the evolving epidemiology of omicron subvariants. Further, the interpretation of the cumulative Covid-19 event curve data is limited by the low numbers of participants at longer follow-up times. Additionally, the trial population was limited to participants ≥16 years of age in the UK who were predominantly white and did not include other key groups such as immunocompromised individuals. Evaluation of longer-term safety of the variant-targeting boosters and the durability of the immune response is ongoing in the study.

In conclusion, the omicron BA.1 bivalent booster mRNA-1273.214 elicited superior neutralizing antibody responses against omicron BA.1 with numerically lower incidences of Covid-19 compared to the original booster vaccine mRNA-1273 in a head-to-head comparison of the two vaccines. Given the continuous and rapid emergence of SARS-CoV-2 variants, it remains important to continuously monitor the neutralization ability as well as vaccine effectiveness against divergent variants.

## Supporting information

Consort checklist

## Data Availability

As the trial is ongoing, access to patient-level data and supporting clinical documents by qualified external researchers may be available upon request and subject to review once the trial is complete.

## Acknowledgments

We thank the study participants and the team of investigators, staff, and colleagues involved in this study for their contributions and dedication; members of the Data Safety and Monitoring Board (Drs. Margaret Johnson [chair], Katrina Pollock, and Chrissie Jones). We thank PPD laboratories for performing neutralization assays and Meso Scale Discovery multiplex binding antibody assays, and Stephanie Lussier for statistical support. Editorial assistance with an early version of the manuscript was provided by Emily Stackpole, Ph.D., and Shane Walton, Ph.D., of MEDiSTRAVA in accordance with Good Publication Practice (GPP3) guidelines, funded by Moderna, Inc., and under the direction of the authors.

