## Supplement for "A Randomized Trial Comparing Omicron-Containing Boosters with the Original Covid-19 Vaccine mRNA-1273"

### Supplementary Appendix

### List of Study Investigators

| Investigator | Institution | Location |
|---|---|---|
| Claire Bethune | University Hospitals Plymouth NHS Trust | Plymouth, United Kingdom |
| Marta Boffito | Chelsea and Westminster Hospital NHS Foundation Trust and Imperial College London | London, United Kingdom |
| Duncan Browne | Royal Cornwall Hospitals NHS Trust | Truro, United Kingdom |
| Marcin Bula | University of Liverpool | Liverpool, United Kingdom |
| Fiona Burns | Royal Free London NHS Foundation Trust University and University College London | London, United Kingdom |
| David Chadwick | South Tees Hospitals NHS Foundation Trust, James Cook University Hospital | Middlesbrough, United Kingdom |
| Rebecca Clark | Layton Medical Centre | Blackpool, United Kingdom |
| Catherine A. Cosgrove (Lead Investigator) | Vaccine Institute, Centre for Neonatal and Paediatric Infection, St George's University of London | London, United Kingdom |
| Paul Dargan | Guy's & St Thomas' NHS Foundation Trust, King's College London | London, United Kingdom |
| Thomas C. Darton | Department of Infection, Immunity and Cardiovascular Disease, University of Sheffield | Sheffield, United Kingdom |
| Christopher J.A. Duncan | Translational and Clinical Research Institute, Newcastle University and NIHR Newcastle Clinical Research Facility, The Newcastle upon Tyne Hospitals NHS Foundation Trust | Newcastle upon Tyne, United Kingdom |
| Stevan Emmett | Royal United Hospitals, Bath NHS Foundation Trust | Bath, United Kingdom |
| James Galloway | Kings College Hospital | London, United Kingdom |
| Lucy Jones | Cwm Taf Morgannwg University Health Board | Wales, United Kingdom |
| Philip A. Kalra | Salford Royal Hospital, Northern Care Alliance NHS Foundation Trust | Salford, United Kingdom |
| Rachel Kaminski | Gloucestershire Hospitals NHS Foundation Trust | Gloucester, United Kingdom |
| Rajeka Lazarus | University Hospitals Bristol and Weston NHS Foundation Trust | Bristol, United Kingdom |
| Patrick Lillie | Hull University Teaching Hospitals NHS Trust | Hull, United Kingdom |
| Patrick Moore | University Hospital Southampton NHS FT | Southampton, United Kingdom |
| Patrick Moore | University Hospital Southampton NHS FT; The Adam Practice, Poole, Dorset, UK, and University Hospital Southampton NHS Foundation Trust | Southampton, United Kingdom |
| Ed Moran | Southmead Hospital | Bristol, United Kingdom |
| Rhiannon Nally | Wansford and Kings Cliffe Practice | Wansford, United Kingdom |
| Adrian Palfreeman | University Hospitals of Leicester NHS Trust, Leicester, UK | Leicester, United Kingdom |
| Tommy Rampling | UCLH Clinical Rese NIHR UCLH Clinical Research Facility and NIHR UCLH Biomedical Research Centre, University College London Hospitals NHS Foundation Trust | London, United Kingdom |
| Anju Sahdev | Barts Health NHS Trust | London, United Kingdom |
| Dinesh Saralaya | National Institute for Health Research Patient Recruitment Centre and Bradford Teaching Hospitals NHS Foundation Trust | Bradford, United Kingdom |
| Ray Sheridan | Royal Devon University Hospital | Exeter, United Kingdom |
| Roy Soiza | Aberdeen Royal Infirmary (NHS Grampian), University of Aberdeen | Aberdeen, Scotland, United Kingdom |

**Methods**

*Trial design*

This study is a phase 2/3, two-part, randomized, observer-blind, active-controlled, multicenter study to evaluate the immunogenicity and safety of mRNA-1273.529 and mRNA-1273.214 booster vaccine in medically stable individuals 16 years and older.
The study consists of 2 parts (Fig. S1). In part 1, the monovalent mRNA-1273.529 comprised of an omicron variant BA.1 (B.1.1.529) mRNA is compared with the original mRNA-1273 booster vaccine, and in part 2, the omicron-containing bivalent mRNA-1273.214 consist of two mRNAs, the S-2P of the SARS-CoV-2 Wuhan Hu-1 strain and the S-2P of the SARS-CoV-2 omicron BA.1 variant.

All eligible participants will have previously received 2 or 3 doses of an authorized/approved Covid-19 vaccine. Participants who had previously received 2 injections of a Covid-19 vaccine as a primary series were to receive mRNA-1273.529, mRNA-1273.214, or mRNA-1273 as the first booster (3rd dose), and participants who previously received a primary series and one booster dose are to receive mRNA-1273.529, mRNA-1273.214, or mRNA-1273 as the second booster (4$^{th}$ dose). Participants who will receive the 4th dose as part of the study must have previously received an mRNA vaccine as the 3rd dose of a Covid-19 vaccine. Participants who will receive the 3rd dose as part of the study may have previously received 2 doses of a mRNA or a non-mRNA Covid-19 vaccine (a mixed regimen approach is acceptable).

*Inclusion and Exclusion Criteria*

Participants were eligible for Part 1 or Part 2 based on the following inclusion criteria:

1. Was male or female, at least 16 years of age at the time of consent
2. Investigator's assessment that the participant understood and was willing and physically able to comply with protocol-mandated follow-up, including all procedures
3. Participant provided written informed consent for participation in this study, including all evaluations and procedures as specified in the protocol
4. Female participants of nonchildbearing potential were enrolled in the study. Nonchildbearing potential was defined as postmenopausal or permanently sterilized; a follicle stimulating hormone level could be measured at the discretion of the investigator to confirm postmenopausal status, if necessary
5. Female participants of childbearing potential were enrolled in the study if the participant fulfilled all the following criteria:
    a. Had a negative pregnancy test at screening and on the day of vaccination prior to vaccine dose being administered on day 1
    b. Had practiced adequate contraception or abstained from all activities that could result in pregnancy for at least 28 days prior to the first dose (day 1); adequate female contraception was defined as consistent and correct use of a local health authority approved contraceptive method in accordance with the product label
    c. Had agreed to continue adequate contraception through 90 days following vaccine administration
6. Had received 2 prior doses of one of the following approved/authorized Covid-19 vaccines: Moderna, Pfizer/BioNTech, Oxford/AstraZeneca, Janssen. A heterologous vaccine regimen was acceptable.

7. Participants who received the fourth dose as part of the study must have previously received a mRNA vaccine (Moderna or Pfizer/BioNTech) as the third dose of a Covid-19 vaccine. Participants who received the third dose as part of the study may have previously received 2 doses of an approved/authorized mRNA or a non-mRNA Covid-19 vaccine (a heterologous vaccine regimen is acceptable).

Participants were excluded from Part 1 or Part 2 based on the following exclusion criteria:

1. Had close contact (without personal protective equipment) as defined by the Centers for Disease Control and Prevention (CDC) in the past 14 days to someone diagnosed with SARS-CoV-2 infection or Covid-19 within 10 days of the close contact. Participants could be rescreened after 14 days provided that they remain asymptomatic
2. Participant was acutely ill or febrile (temperature ≥38.0°C/100.4°F) 72 hours prior to or at the Screening Visit or day 1. Participants meeting this criterion could be rescheduled within the 28-day screening window and will retain their initially assigned participant number
3. Had tested positive for SARS-CoV-2 by an authorized/approved lateral flow/rapid antigen or PCR test within 90 days of Screening
4. Had received a Covid-19 vaccine within 90 days of the Screening Visit
5. Had received a total of 4 doses or more of Covid-19 vaccine
6. Had received a Covid-19 vaccine at a dose different from the authorized/approved dose.
7. History of a diagnosis or condition that, in the judgment of the Investigator, was clinically unstable or may affect participant safety, assessment of safety endpoints, assessment of immune response, or adherence to study procedures. Clinically unstable was defined as a diagnosis or condition requiring significant changes in management or medication within the 2 months prior to screening and includes ongoing workup of an undiagnosed illness that could lead to a new diagnosis or condition
8. Reported history of congenital or acquired immunodeficiency, immunosuppressive condition, or immune-mediated disease requiring immunosuppressive treatment or other immunosuppressive condition
9. Dermatologic conditions that could affect local solicited AR assessments (eg, tattoos, psoriasis patches affecting skin over the deltoid areas)
10. Reported history of anaphylaxis or severe hypersensitivity reaction after receipt of any components of mRNA vaccine
11. Reported history of bleeding disorder that is considered a contraindication to intramuscular injection or phlebotomy
12. Any medical, psychiatric, or occupational condition, including reported history of substance abuse, that, in the opinion of the Investigator, might pose additional risk due to participation in the study or could interfere with the interpretation of study results
13. Had received systemic immunosuppressants or immune-modifying drugs for >14 days in total within 181 days prior to screening (for corticosteroids ≥10 mg/day of prednisone or equivalent) or was anticipating the need for immunosuppressive treatment at any time during participation in the study
14. Had received or planned to receive any licensed vaccine ≤28 days prior to the study injection (day 1) or planned to receive a licensed vaccine within 28 days after the study injection (with the exception that approved seasonal influenza vaccine may be received by at least 7 days and preferably 14 days apart from the study injection)

15. Had received systemic immunoglobulins or blood products within 90 days prior to the Screening Visit or plans to receive during the study
16. Diagnosis of malignancy within the previous 10 years (excluding nonmelanoma skin cancer)
17. Had donated ≥ 450 mL of blood products within 28 days prior to the Screening Visit or planned to donate blood products during the study
18. Had participated in an interventional clinical study within 28 days prior to the Screening Visit based on the medical history interview or planned to do so while participating in this study
19. Was an immediate family or household member of study personnel, study site staff, or Sponsor personnel

*Trial Vaccine*
The monovalent mRNA-1273.529 vaccine contains mRNA encoding for the S-2P of the SARS-CoV-2 omicron variant BA.1 (B.1.1.529). The mRNA-1273.214 vaccine contains two mRNAs encoding for both the S-2P of the SARS-CoV-2 Wuhan Hu-1 strain and the S-2P of the SARS-CoV-2 omicron BA.1 variant, formulated in the same LNP.

*Randomization and Masking*
Randomization during both parts was performed using an interactive response technology. In part 1, approximately 600-1000 participants were planned to be randomized in a 1:1 ratio to receive a single dose of either 50 µg of mRNA-1273.529 or 50 µg of mRNA-1273 (active control). In part 2, approximately 2,924 participants were planned to be randomized in a 1:1 ratio to receive a single dose of either 50 µg of mRNA-1273.214 or 50 µg of mRNA-1273 (active control). In both parts, randomization was stratified by age groups (16 to <65 years or ≥65 years) and number of booster doses received (to receive study vaccine as the 4th dose or to receive study vaccine as the 3rd dose). At least ≥90% of participants were prespecified to receive study vaccine as the 4th dose.

Enrollment was observer-blinded to treatment assignment because the study vaccines differed in appearance. Dose preparation, administration, and accountability was performed by designated site personnel who did not participate in any clinical study evaluations. The unblinded site personnel prepared the dose out of view of the participant as well as blinded site personnel, and did not reveal the identity of the study vaccine except in case of emergency. Laboratory personnel responsible for immunogenicity testing were blinded to the treatment assignment of the samples tested throughout the study.

*Sample Size Determination*
For sample size determination in Part 1, approximately 300 to 500 participants per vaccine group were targeted for enrollment (200 to 337 evaluable participants for each vaccine group) to provide >95% power to demonstrate noninferiority of mRNA-1273.529 against omicron BA.1 at 2-sided α of 1.0% at day 29. With this sample size range, the power to demonstrate superiority of mRNA-1273.529 against omicron BA.1 at a 2-sided α of 1.0% was 14% to 82% at day 29. Assumptions included that the true GMC ratio (mRNA-1273.529 versus mRNA-1273) ranged from 1.25 to 1.5, the standard deviation of the natural log-transformed concentration was 1.5, and a non-inferiority margin of 1.5. In Part 2, the target enrollment was 1328 participants for each vaccine group among those who received the study vaccine as a fourth

dose. Assuming 996 evaluable participants in each arm, there was more than 95% power to demonstrate noninferiority of mRNA-1273.214 against omicron BA.1 and against the ancestral SARS-CoV-2 (D614G) at 2-sided α of 1.0% at day 29; this sample size would also provide 77% to >95% power to demonstrate superiority of mRNA-1273.214 against omicron BA.1 at a 2-sided α of 1.0% at day 29. Assumptions included that the true GMC ratio (mRNA-1273.214 versus mRNA-1273) ranged from 1.25 to 1.5 for omicron BA.1 and was 1 for the ancestral SARS-CoV-2 (D614G), the standard deviation of the natural log-transformed concentration was 1.5, and a non-inferiority margin of 1.5.

*Immunogenicity Statistical Analyses*

*Statistical hypotheses*

Part 1:

The primary objective on immune response in part 1 of this study is based on the participants who will receive the second boosters (4th dose). There are two identical hypotheses, one each for day 29 and month 3 timepoints.

Primary Hypotheses:

1) mRNA-1273.529, as a single booster dose, is non-inferior to mRNA-1273 based on GMC ratio against the BA.1 strain with a non-inferiority margin of 1.5 at day 29.

2) mRNA-1273.529, as a single booster dose, is non-inferior to mRNA-1273 based on GMC ratio against the BA.1 strain with a non-inferiority margin of 1.5 at Month 3.

Key Secondary Hypothesis:

mRNA-1273.529, as a single booster dose, is superior to mRNA-1273 based on GMC ratio against the BA.1 strain at day 29 or Month 3.

For the primary objective of immune response, hypotheses testing based on participants receiving the booster (4th dose), alpha of 0.05 (2-sided) were allocated to the 2 time points: alpha of 0.01 (2-sided) was allocated to day 29, alpha of 0.04 (2-sided) was allocated to month 3. Only interim analysis results for the day 29 hypotheses are presented in this report; hypotheses for month 3 will be tested later when month 3 data become available.

The non-inferiority of mRNA-1273.529 as compared to mRNA-1273 against the BA.1 strain at day 29 was assessed using a non-inferiority margin of 1.5 at 2-sided alpha of 0.01. The primary immunogenicity objective is considered met if non-inferiority against the BA.1 strain is demonstrated, ie, the lower bound of the 99% confidence interval (CI) of the GMC ratio of mRNA-1273.529 vs. mRNA-1273 against BA.1 is ≥ 0.67 (non-inferiority margin of 1.5).

The non-inferiority of mRNA-1273.529 as compared to mRNA-1273 against the BA.1 strain at month 3 will be assessed using a non-inferiority margin of 1.5 at 2-sided alpha of 0.04. The primary immunogenicity objective is considered met if non-inferiority against the BA.1 strain is demonstrated, ie, the lower bound of the 96% confidence interval (CI) of the GMC ratio of mRNA-1273.529 vs. mRNA-1273 against BA.1 is ≥0.67 (non-inferiority margin of 1.5).

Part 2:

The primary objective on immune response in part 2 of this study is based on the participants who received second booster doses (4th dose) in part 2. There are 6 hypotheses on immune response of mRNA-1273.214 listed below with 3 identical hypotheses for day 29 and month 3. Part 2 of this study is considered to have met its primary objective if non-inferiority of mRNA-1273.214 against the BA.1 strain, non-inferiority of mRNA- 1273.214 against the prototype strain, and superiority of mRNA-1273.214 against the BA.1 strain are demonstrated as compared to mRNA-1273 at day 29 or month 3. Only interim analysis results for the day 29 hypotheses are presented in this report; hypotheses for month 3 will be tested later when month 3 data become available. Fig. S3 depicts the hypotheses testing strategy.

Primary Hypotheses:

1) mRNA-1273.214, as a single booster dose, is non-inferior to mRNA-1273 based on GMC ratio against the BA.1 strain with a non-inferiority margin of 1.5 at day 29.

2) mRNA-1273.214, as a single booster dose, is non-inferior to mRNA-1273 based on GMC ratio against the prototype strain with a non-inferiority margin of 1.5 at day 29.

3) mRNA-1273.214, as a single booster dose, is superior to mRNA-1273 based on GMC ratio against the BA.1 strain at day 29.

4) mRNA-1273.214, as a single booster dose, is non-inferior to mRNA-1273 based on GMC ratio against the BA.1 strain with a non-inferiority margin of 1.5 at Month 3.

5) mRNA 1273.214, as a single booster dose, is non-inferior to mRNA-1273 based on GMC ratio against the prototype strain with a non-inferiority margin of 1.5 at Month 3.

6) mRNA-1273.214, as a single booster dose, is superior to mRNA-1273 based on GMC ratio against the BA.1 strain at Month 3.

For the part 2 primary objective of immune response, hypothesis testing based on participants receiving the second booster dose (4th dose), alpha of 0.05 (2-sided) were allocated to the 2 timepoints: alpha of 0.01 (2-sided) was allocated to day 29, and alpha of 0.04 (2-sided) was allocated to month 3 (Fig. S3). The non-inferiority of mRNA-1273.214 as compared to mRNA-1273 against the BA.1 strain and the non-inferiority of mRNA-1273.214 as compared to mRNA-1273 against the prototype strain at day 29 are assessed using a non-inferiority margin of 1.5 at 2-sided alpha of 0.01. The primary immunogenicity objective is considered met if non-inferiority against the BA.1 strain and the ancestral SARS-CoV-2 (D614G) are both demonstrated, ie, the lower bound of the 99% confidence interval (CI) of the GMC ratio of mRNA-1273.214 vs. mRNA-1273 against BA.1 is ≥0.67 (non-inferiority margin of 1.5) and the lower bound of the 99% CI of the GMR of mRNA-1273.214 vs mRNA-1273 against prototype is ≥0.67. Once the non-inferiority of mRNA-1273.214 as compared to mRNA-1273 against the BA.1 strain and against ancestral SARS-CoV-2 (D614G) is demonstrated, the superiority of mRNA-1273.214 as compared to mRNA-1273 against the BA.1 strain is evaluated at day 29. The 99% CI of GMR (mRNA-1273.214 vs. mRNA-1273) is used to assess superiority and if the lower bound of the GMR rules out (>1) at day 29, superiority of mRNA-1273.214 compared to mRNA-1273 against BA.1 is considered demonstrated.

In part 1, an analysis of covariance (ANCOVA) model was used to assess the difference in immune response between mRNA-1273.529 and mRNA-1273 in participants who received the study vaccine as their fourth dose. In this model, antibody concentrations against omicron BA.1 at day 29 after vaccination was a dependent variable, and a group variable (mRNA-1273.529 and mRNA-1273) was the fixed effect, while adjusting for age groups (aged <65 or ≥65 years), most recent Covid-19 vaccination type (mRNA, viral vector), and pre-booster antibody concentration. The GMC was estimated by the geometric least square mean (GLSM) from the model and its corresponding 99% confidence interval (CIs) were calculated for each vaccine group. The GMR for mRNA-1273.529 versus mRNA-1273 was estimated by the ratio of GLSM from the model, with corresponding 99% CIs determined. The 99% CI for the GMR was used for non-inferiority testing to assess the between group difference (mRNA-1273.529 versus mRNA-1273) in immune response against omicron BA.1 at day 29. Non-inferiority of immune response against omicron BA.1 was declared if the lower bound of the corresponding 99% CI was ≥0.67 based on a non-inferiority margin of 1.5. Dependent on if non-inferiority was demonstrated, superiority of mRNA-1273.529 versus mRNA-1273 against omicron BA.1 was subsequently tested; superiority was considered demonstrated if the lower bound of the 99% CI of the GMC ratio was >1 at day 29. Similar methodologies were used to assess immune responses against ancestral SARS-CoV-2 (D614G). The SRR is measured by an increase of SARS-CoV-2–specific bAb or GMC from pre-booster below the lower limit of quantification (< LLOQ) to at least 4 x LLOQ, or a 4-fold or greater rise if pre-booster ≥ LLOQ. The SRR is summarized at day 29 for each treatment group with the 95% CI calculated using the Clopper-Pearson method. The difference of SRRs at all day 29 for mRNA-1273.529 compared with mRNA-1273 will be provided with 95% CI using Miettinen-Nurminen method.

In part 2, an ANCOVA model was used to assess the difference in immune response between mRNA-1273.214 and mRNA-1273 in the participants who received the study vaccine as the fourth dose. Similar to analyses performed in part 1, antibody concentrations against omicron BA.1 and ancestral SARS-CoV-2 (D614G) at day 29 after vaccination was a dependent variable and a group variable (mRNA-1273.214 and mRNA-1273) was the fixed effect, with adjustments for age groups (aged <65 or ≥65 years), most recent Covid-19 vaccination type (mRNA, viral vector), and pre-booster antibody concentration. The GMC was estimated by the GLSM from the model, with corresponding 99% CIs provided for each vaccine group and the GMR for mRNA-1273.214 compared with mRNA-1273 was estimated by the ratio of GLSM. The corresponding 99% CIs for the GMR were calculated and used to assess the difference in immune response for mRNA-1273.214 compared to mRNA-1273 at day 29 for non-inferiority of omicron BA.1 and ancestral SARS-CoV-2 (D614G) and superiority of omicron BA.1 testing. This same analysis was used for secondary analyses on immune responses against other SARS-CoV-2 variants with a 95% CI for the GMR used for non-inferiority and superiority. The SRR is measured by an increase of SARS-CoV-2–specific bAb or GMC from pre-booster below the lower limit of quantification (< LLOQ) to at least 4 x LLOQ, or a 4-fold or greater rise if pre-booster ≥ LLOQ. The SRR is summarized at day 29 for each treatment group with the 95% CI calculated using the Clopper-Pearson method. The difference of SRRs at all day 29 for mRNA-1273.214 compared with mRNA-1273 will be provided with 95% CI using Miettinen-Nurminen method.

Additional immunogenicity analysis included observed nAb GMC in all participants regardless of SARS-CoV-2-infection status and among those with evidence of prior SARS-CoV-2-infection in part 2 and observed bAb GMCs and GMRs (95% CI) against variants assessed by ANCOVA in parts 2 are also provided.

*Immunogenicity Assays*

*Pseudovirus neutralization assays*

For the assessment of ancestral SARS-CoV-2 (D614G) neutralizing antibodies, post-vaccination serology samples from participants were tested using a validated reporter virus microneutralization assay (PPD, part of Thermo Fisher Scientific Vaccines Laboratory Services, Richmond, Virginia).[1,2] This cell-based assay measures the SARS-CoV-2 neutralizing antibody inhibition of the infection of 293T-ACE2 cells by SARS-CoV-2 reporter virus particles (RVP: Wuhan-Hu-1 isolate including D614G). The serum antibody concentration is determined by interpolating the mean of the replicate foci forming unit values from the fitted reference standard curve calibrated to the first WHO International Antibody Standard for SARS-CoV-2 Lot 20/136. The interpolated antibody concentrations were dilution corrected, and the final concentration is the antibody concentration associated with the lowest dilution with an antibody concentration within the quantifiable range of the assay. The results are reported as final antibody geometric mean concentration (GMC) in AU/mL.

For the assessment of Omicron BA.1 neutralizing antibodies, post-vaccination serology samples from participants were tested using a validated reporter virus microneutralization assay (PPD, part of Thermo Fisher Scientific Vaccines Laboratory Services, Richmond, Virginia). The assay methodology is the same as the above-mentioned neutralization assay while utilizing SARS-CoV-2 reporter virus particles containing mutations of the spike (omicron BA.1) variant.

*Meso Scale Discovery (MSD) binding antibody assay*

The validated Meso Scale Discovery (MSD, Rockville, MD) assay (SARSCOV2S2P [VAC123]; was used for the detection of binding antibody against SARS-CoV2 variants.[3] The assay uses an indirect, quantitative, electrochemiluminescence method to detect SARS-CoV-2 binding IgG antibodies that bind to the SARS-CoV-2 full-length spike protein (Wuhan-Hu-1 ancestral SARS-CoV-2; alpha [B.1.1.7] with the following amino acid changes in the spike protein [ΔH69-V70, ΔY144, N501Y, A570D, D614G, P681H, T761I, S982A, and D1118H]; gamma [P.1] with the following amino acid changes in the spike protein [L18F, T20N, P26S, D138Y, R190S, K417T, E484K, N501Y, D614G, H655Y, T1027I, and V1176F]); delta [B.1.617.2; AY.4; Alt Seq 2] with the following amino acid changes in the spike protein [T19R, T95I, G142D, Δ156/157, R158G, L452R, T478K, D614G, P681R, and D950N]); omicron [B.1.1.529; BA.1] with the following amino acid changes in the spike protein [A67V, ΔH69-V70, T95I, G142D, Δ143-145, Δ211/L212I, ins214EPE, G339D, S371L, S373P, S375F, K417N, N440K, G446S, S477N, T478K, E484A, Q493R, G496S, Q498R, N501Y, Y505H, T547K, D614G, H655Y, N679K, P681H, N764K, D796Y, N856K, Q954H, N969K, and L981F]) in human serum. The assay was performed by PPD Vaccine Laboratory, Richmond VA, and is based on MSD technology which employs capture molecule MULTI-SPOT® microtiter plates fitted with a series of electrodes.

*<u>Incidence of SARS-CoV-2 infection</u>*

*Assessments for SARS-CoV-2 Infection*

Participants were directed as soon as possible and within 24 hours to obtain an approved/authorized PCR test for SARS-CoV-2 locally outside of the study according to prior National Health Service (NHS) guidance if they experience symptoms of Covid-19 as previously defined by the NHS:

- High temperature (feel hot to touch on the chest or back)
- A new, continuous cough (coughing a lot for more than an hour, or 3 or more coughing episodes in 24 hours, or worse than usual cough)
- A loss or change to sense of smell or taste

If a PCR test was unavailable locally, participants were requested to come in for a study visit or take an approved/authorized lateral flow/rapid antigen test for SARS-CoV-2. Participants were directed as soon as possible and within 24 hours to take an approved/authorized lateral flow/rapid antigen test for SARS-CoV-2 if they experienced any of the following (but did not meet the symptoms of Covid-19 as previously defined by the NHS and described above):

- Signs or symptoms of SARS-CoV-2 infection as defined by the CDC with modifications:
    - Chills
    - Cough (not meeting Covid-19 symptoms defined by the NHS)
    - Shortness of breath or difficulty breathing

- Fatigue
- Muscle or body aches
- Headache
- Sore throat
- Congestion or runny nose
- Nausea or vomiting
- Diarrhea

Lateral flow/rapid antigen test for SARS-CoV-2 could be repeated every 24-48 hours if participants continued to have symptoms above.

Participants were directed to take an approved/authorized lateral flow/rapid antigen test for SARS-CoV-2 within 4 to 6 days after last exposure* (see definition of last exposure below) if a participant experienced the following:

- Known close contact with someone who has known Covid-19 or SARS-CoV-2 infection. Examples include:
    - Being within 2 meters (without personal protective equipment (PPE)) for a total of 15 minutes or more
    - Providing care at home
    - Having direct physical contact (hugged or kissed them)
    - Sharing eating or drinking utensils
    - Being sneezed or coughed upon or getting respiratory droplets on the participant

*Last exposure was defined as the last day a participant was in close contact with a symptomatic person in the household (if the participant was then isolated from that person) or the last day of the quarantine of the person the participant was exposed to, if that person was asymptomatic and/or the participant was unable to isolate from that person.

A study illness visit (study site visit) was arranged as soon as possible and within 72 hours for participants who test positive or equivocal for SARS-CoV-2 using an authorized/approved lateral flow/rapid antigen or local PCR testing. If a test for SARS-CoV-2 was unavailable or if there was uncertainty on the test result, a study site visit may be arranged as soon as possible and within 72 hours. At this visit, an NP swab was collected to evaluate the presence of SARS-CoV-2 infection. Active surveillance for Covid-19 was also conducted throughout the study according to the schedule of events.

The incidence of symptomatic and asymptomatic SARS-CoV-2 infection is an exploratory endpoint in part 1 and a secondary endpoint in part 2, assessed 14 days after randomization.

*SARS-CoV-2 infection*

SARS-CoV-2 infection is defined in participants with negative SARS-CoV-2 status pre-booster on the basis of either binding antibody (bAb) levels against SARS-CoV-2 nucleocapsid protein negative (as measured by Roche Elecsys) at day 1 that becomes positive (as measured by Roche Elecsys) post-baseline, OR positive RT-PCR post-baseline. The date of documented infection will be the earlier of the date of positive post-baseline RT-PCR result, or date of a positive serology test result based on bAb specific to SARS-CoV-2 nucleocapsid. Cases are counted starting 14 days after the randomization (date of documented infection – date of randomization ≥14). The time to the first SARS-CoV-2 infection is calculated as the time to 1st SARS-CoV-2 infection (date of the 1st documented infection – date of randomization + 1). The incidence of SARS-CoV-2 infection counted starting 14 days after randomization is summarized by treatment group. Supportive analyses summarize incidence of SARS-CoV-2 infection in which a case is counted after randomization, i.e., date of documented infection – date of randomization ≥ 0.

*Asymptomatic SARS-CoV-2 infection*

The incidence of asymptomatic SARS-CoV-2 infection is measured by RT-PCR and/or serology tests obtained at post-baseline visits counted starting 14 days after randomization in participants with negative SARS-CoV-2 status pre-booster. Asymptomatic SARS-CoV-2 infection is identified by absence of symptoms and infections as detected by RT-PCR or serology tests. Specifically, the absence of Covid-19 symptoms +/- 14 days AND at least one bAb level against SARS-CoV-2 nucleocapsid protein negative (as measured by Roche Elecsys) at day 1 that becomes positive (as measured by Roche Elecsys) post-baseline, or positive RT-PCR test post-baseline (at scheduled or unscheduled/illness visits). The date of documented asymptomatic infection is the earlier date of positive serology test result based on bAb specific to SARS-CoV-2 nucleocapsid due to infection, or positive RT-PCR at scheduled visits, with the absence of symptoms. The time to the asymptomatic SARS-CoV-2 infection is calculated as the time to the asymptomatic SARS-CoV-2 infection (date of asymptomatic SARS-CoV-2 infection – date of randomization + 1).

*Symptomatic SARS-CoV-2 Infection (Covid-19)*

The incidence of symptomatic SARS-CoV-2 infection starting 14 days after randomization is an exploratory endpoint in part 1 and a secondary endpoint in part 2. Participants had a baseline (day 1) evaluation for SARS-CoV-2 infection and ongoing surveillance for Covid-19 throughout the study. Surveillance for Covid-19 symptoms is conducted via safety calls throughout the duration of the study and blood draw from scheduled visits. Participants reporting certain Covid-19 symptoms have an unscheduled visit to collect a nasopharyngeal swab. Symptomatic SARS-CoV-2 infection is defined in the following two ways:

1. Protocol-defined Covid-19 case definition: The participant must have experienced at least TWO of the following systemic symptoms: Fever (≥ 38ºC), chills, myalgia, headache, sore throat, new olfactory and taste disorder(s), OR the participant must have experienced at least ONE of the following respiratory signs/symptoms: cough, shortness of breath or difficulty breathing, OR clinical or radiographical evidence of pneumonia; AND the participant must have at least one nasopharyngeal (NP) swab, nasal swab, or saliva sample (or respiratory sample, if hospitalized) positive for SARS-CoV-2 by RT-PCR
2. CDC Case Definition of Covid-19 (secondary case definition): Cases will be identified as a positive post-baseline RT-PCR test result that is prompted by symptom(s), AND at least ONE of the following systemic symptoms: Fever (temperature ≥ 38ºC/≥ 100.4ºF) or chills, fatigue, muscle and/or body aches (unrelated to exercise), headache, congestion or runny nose, new loss of taste or smell, sore throat, or vomiting or diarrhea; OR at least ONE of the following respiratory signs/symptoms: cough, shortness of breath or difficulty breathing. (c)

The date of documented Covid-19 (case) is the later date of the eligible symptom(s) and date of positive PCR test and the two dates should be within 14 of each other. Specifically, for the primary definition, the date of the documented Covid-19 is the later date of the positive RT-PCR test, eligible symptom(s), defined as earliest of systemic symptoms: earliest date of the 2nd eligible systemic symptom is reported, earliest date of an eligible respiratory symptom is reported.

For the secondary definition of a Covid-19 infection, the date of documented Covid-19

is the later date of positive RT-PCR test, eligible symptom(s), defined as earliest of earliest date of an eligible respiratory symptom is reported, earliest date of the eligible systemic symptom is reported. The time to the first occurrence of Covid-19 is be calculated as:
date of documented Case Definition of Covid-19 – date of randomization + 1.
Cases are counted starting 14 days post randomization, i.e., date of documented Covid-19 – Date of randomization ≥ 14.

### Figure S1. Study design

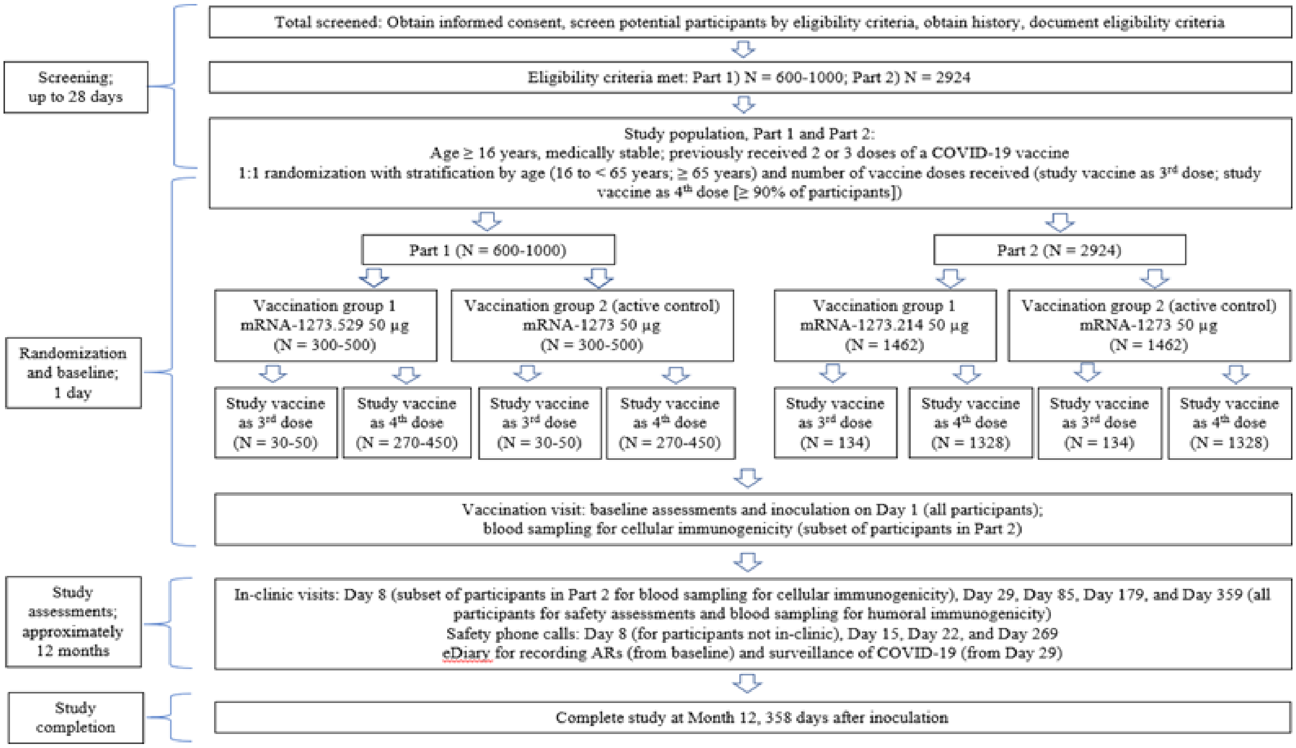


Abbreviations: Covid-19 = coronavirus disease 2019; mRNA = messenger ribonucleic acid; N = number.

#### Figure S2. Analysis populations trial profile

Part 1

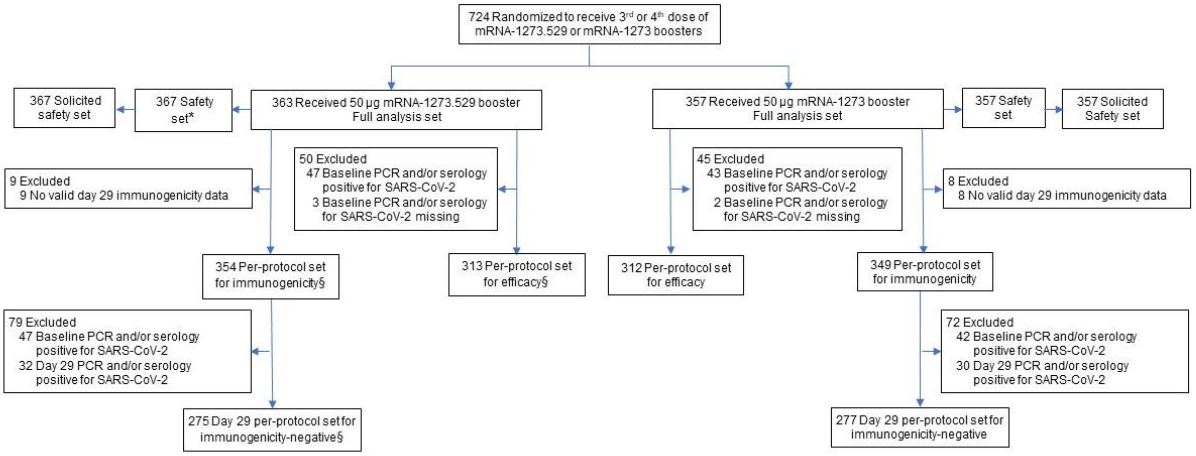


Part 2

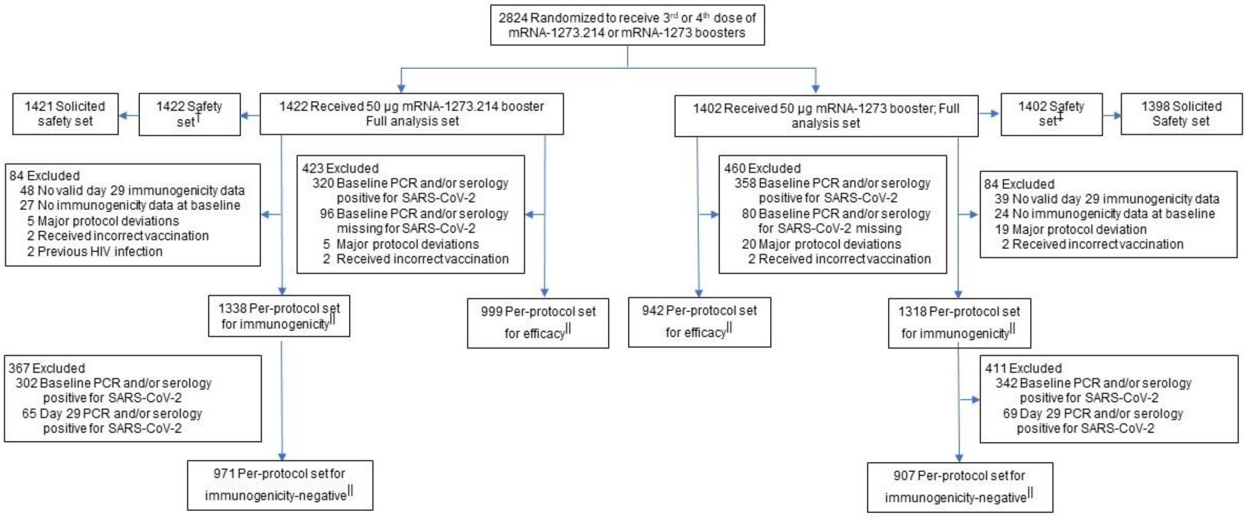


**Figure. S2.** The full analysis set (FAS) consists of all participants who receive study vaccine. The safety set consists of all participants who receive study vaccine (both first [third] and second [fourth] booster doses) and was used for all analyses of safety except for solicited adverse reactions which were assessed in the solicited safety set. The solicited safety set includes participants in the safety set who contribute a solicited adverse reaction. The per-protocol set for immunogenicity consists of all participants in the FAS who received the planned second (fourth) booster dose of study vaccination and had no major protocol deviations that impact key or critical data. The per-protocol set for immunogenicity-negative set consists of participants in the per-protocol set for immunogenicity who have no serologic or virologic evidence of SARS-CoV-2 infection up to the day of visit analysis, i.e., who are SARS-CoV-2 negative, defined by both negative RT-PCR test for SARS-CoV-2 and negative serology test based on bAb specific to SARS-

CoV-2 nucleocapsid and is the primary analysis set for immunogenicity comparisons between boosters s. *The part 1 safety set includes 4 participants who due to a randomization error, were randomized to a non-study arm but received the mRNA-1273.529 50-µg study vaccine and were not included in the FAS; the safety set also includes one participant who received only a third dose of mRNA-1273.529. †The safety set includes four participants who received a third dose of mRNA-1273.214. ‡The safety set includes seven participants who received a third dose of mRNA-1273. §Part 1 includes 1 participant in the per-protocol immunogenicity set, day 29 per-protocol immunogenicity set-negative and per-protocol efficacy set in the mRNA-1273.529 arm who received a third dose only and was excluded from the analyses. ||Part 2 includes 4 and 7 participants in the per-protocol immunogenicity set and 2 and 5 participants in the day 29 per-protocol immunogenicity set-negative and per-protocol efficacy set, respectively in the mRNA-1273.214 and mRNA-1273 arms who received a third dose only and were excluded from the analyses.

**Figure S3. Statistical Testing Sequence for Immunogenicity in Part 2**

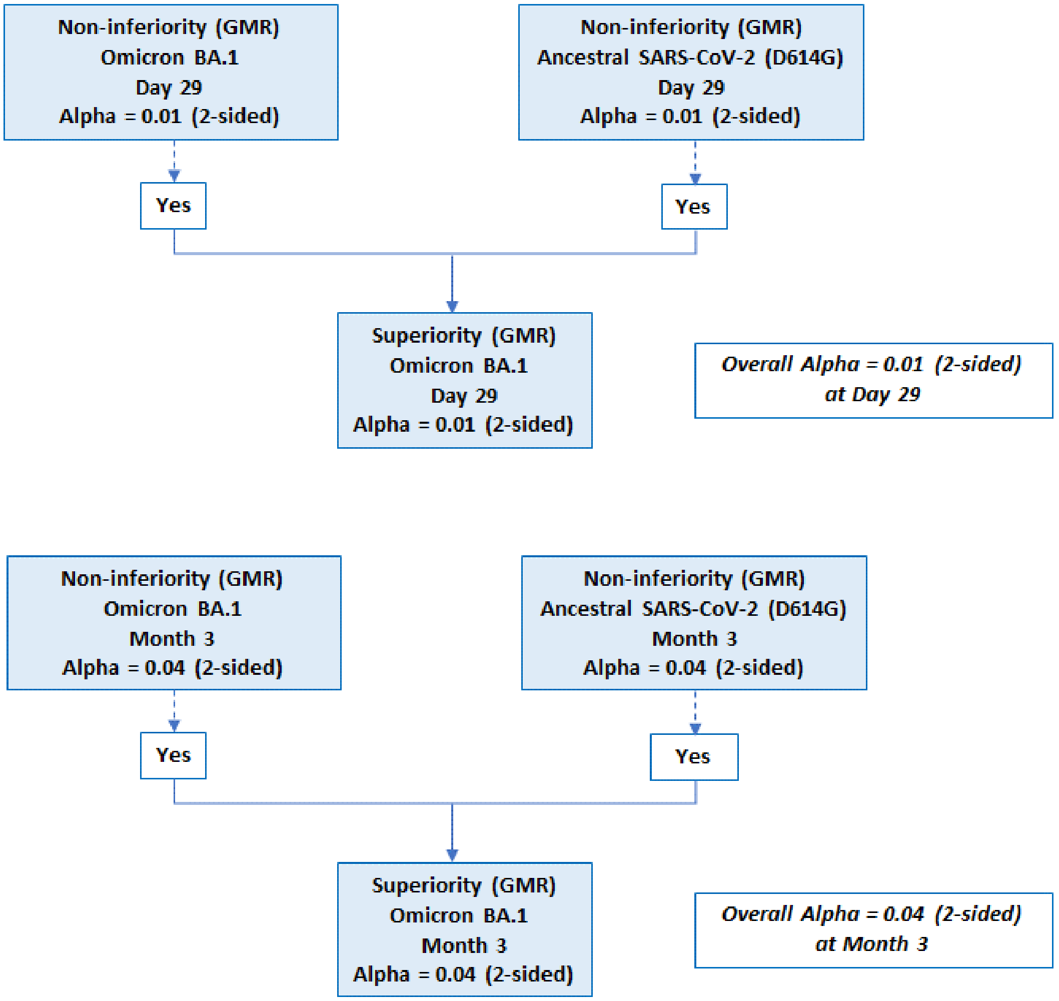


**Figure S3.** GMR=geometric mean concentration ratio. A hierarchical testing approach was used for the primary immunogenicity objectives in part 2 of the study and were tested with the family-wise type 1 alpha of 0.05 (2-sided), allocated to day 29 (alpha 0.01 [2-sided]) and month 3 (alpha 0.04 [2-sided]) timepoints (Fig. S3). At both days, there were 3 prespecified endpoints based on GMR for the primary objectives: 1) non-inferiority of the antibody response of mRNA-1273.214 versus mRNA-1273 against the BA.1 strain, 2) non-inferiority of mRNA-1273.214 versus mRNA-1273 against ancestral SARS-CoV-2 (D614G) and 3) the superiority of mRNA-1273 versus mRNA-1273 against the BA.1 strain (detailed in supplementary methods). The endpoint testing sequence prespecified that both non-inferiority endpoints must first be met before testing for the superiority of the antibody response against omicron BA.1. Only results at day 29 are included in this interim analysis. The two primary immunogenicity objectives are considered met if the lower bound of the 99% CI of the GMR of mRNA-1273.214 versus mRNA-1273 at day against BA.1 and ancestral SARS-CoV-2 (D614G) is ≥0.67 with a non-inferiority margin of 1.5. If the non-inferiority of mRNA-1273.214 versus mRNA-1273 antibody response against the BA.1 strain and ancestral SARS-CoV-2 (D614G) is demonstrated, the superiority of the response of mRNA-1273.214 versus mRNA-1273 against the BA.1 strain is evaluated. Superiority of mRNA-1273.214 compared to mRNA-1273 against BA.1 is considered demonstrated if the lower bound of the 99% CI of the GMR rules out (>1) at day 29.

**Figure S4. Percentages of Prior Vaccinations and Booster Doses in Participants**

**A. Part 1**

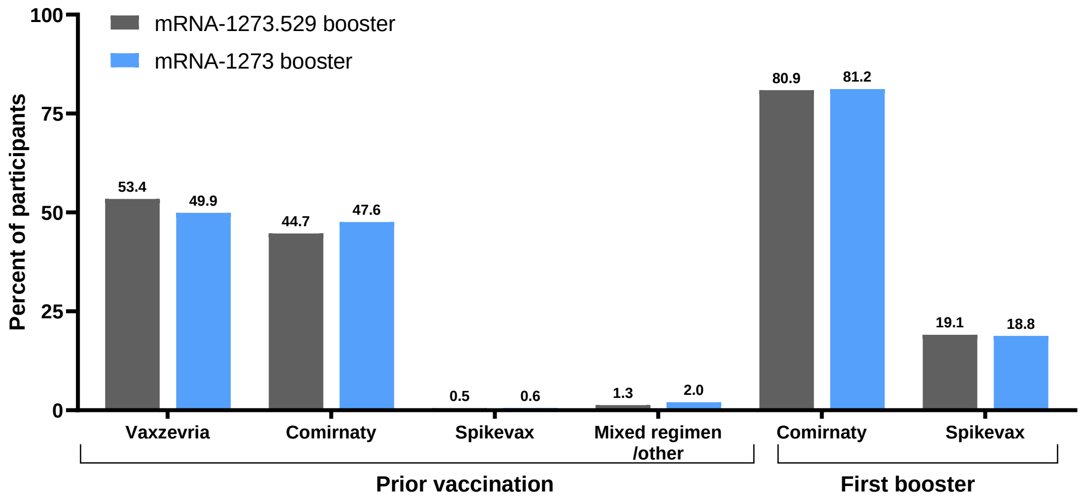


**B. Part 2**

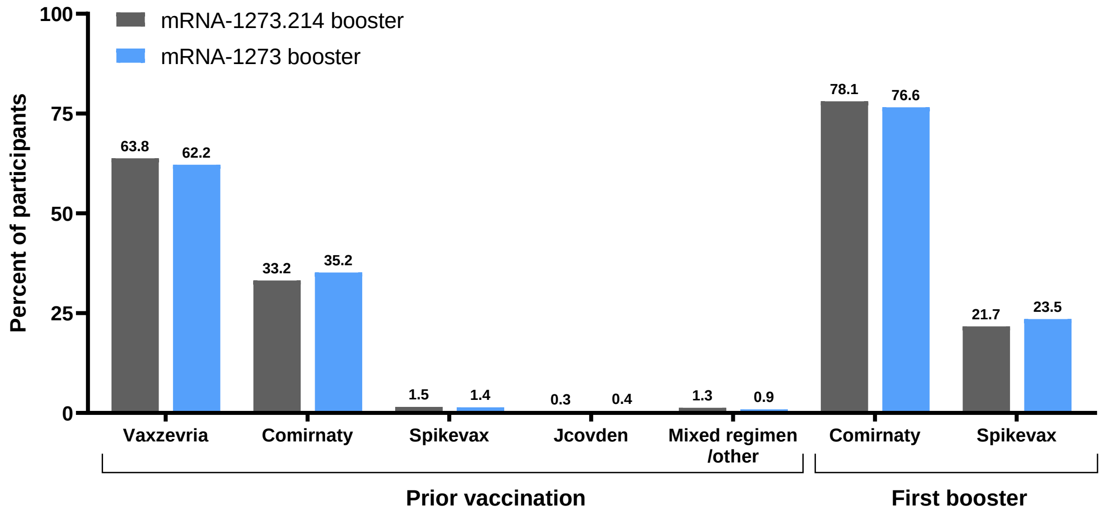

**Figure S5. Observed Pseudovirus Neutralizing Antibodies Against Omicron BA.1 or Ancestral SARS-CoV-2 (D614G) after Receipt of mRNA-1273.529, mRNA-1273.214, or mRNA-1273 Administered as Second Boosters to Participants with No Prior SARS-CoV-2-Infection**

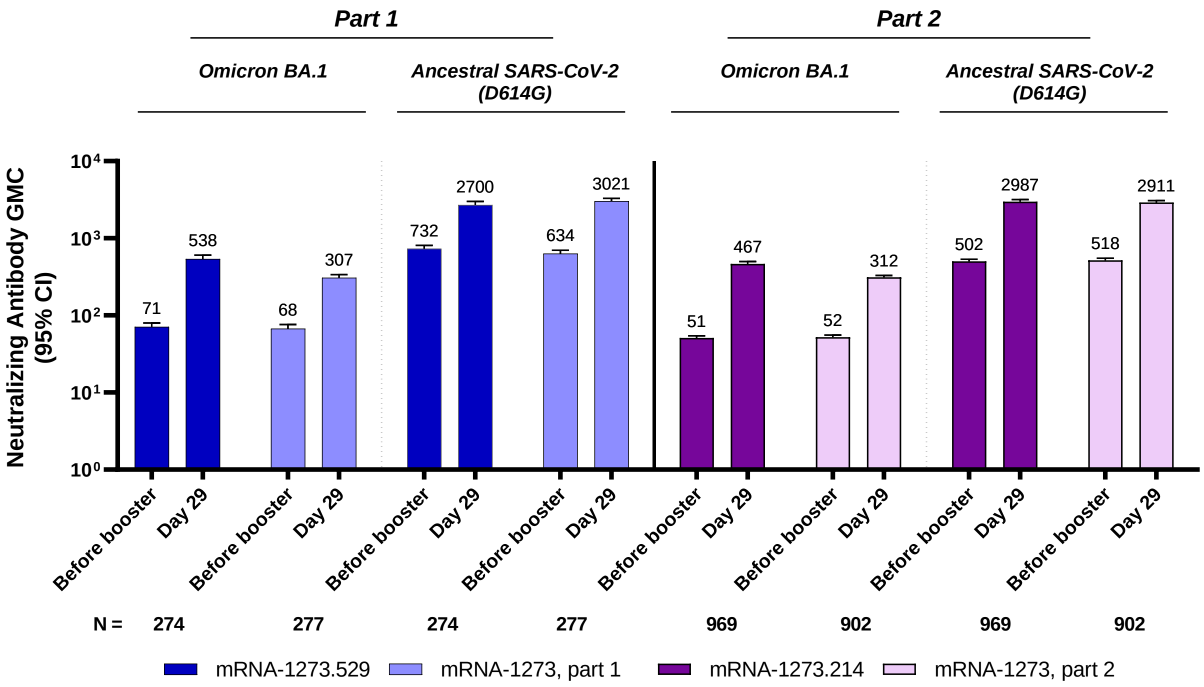


**Figure S5:** Geometric mean concentrations of pseudovirus neutralizing antibodies are shown for the per-protocol set for immunogenicity-negative set (no prior SARS-CoV-2 infection) (Part 1, N=274 in the mRNA-1273.529 and N=277 in the mRNA-1273 groups; Part 2, N=969 in the mRNA-1273.214 and 902 in the mRNA-1273 groups). Antibody values that were reported as being below the lower limit of quantification (LLOQ: 8 for omicron BA.1; 10 for ancestral SARS-CoV-2 [D614G]) were replaced by 0.5 times the LLOQ. Values greater than the upper limit of quantification (ULOQ: 41,984 for omicron BA.1; 4,505,600 for ancestral SARS-CoV-2 [D614G]) were replaced by the ULOQ if actual values were not available. Included are participants with no previous SARS-CoV-2 infection (primary analysis set). The 95% confidence intervals (95% CIs) were calculated based on the t-distribution of the log-transformed values for geometric mean values, then back transformed to the original scale for presentation.

**Figure S6: Observed Pseudovirus Neutralizing Antibodies Against Omicron BA.1 and Ancestral SARS-CoV-2 (D614G) after Receipt of mRNA-1273.214 or mRNA-1273 Administered as Second Boosters by Prior SARS-CoV-2 Infection Pre-Booster Status, Part 2**

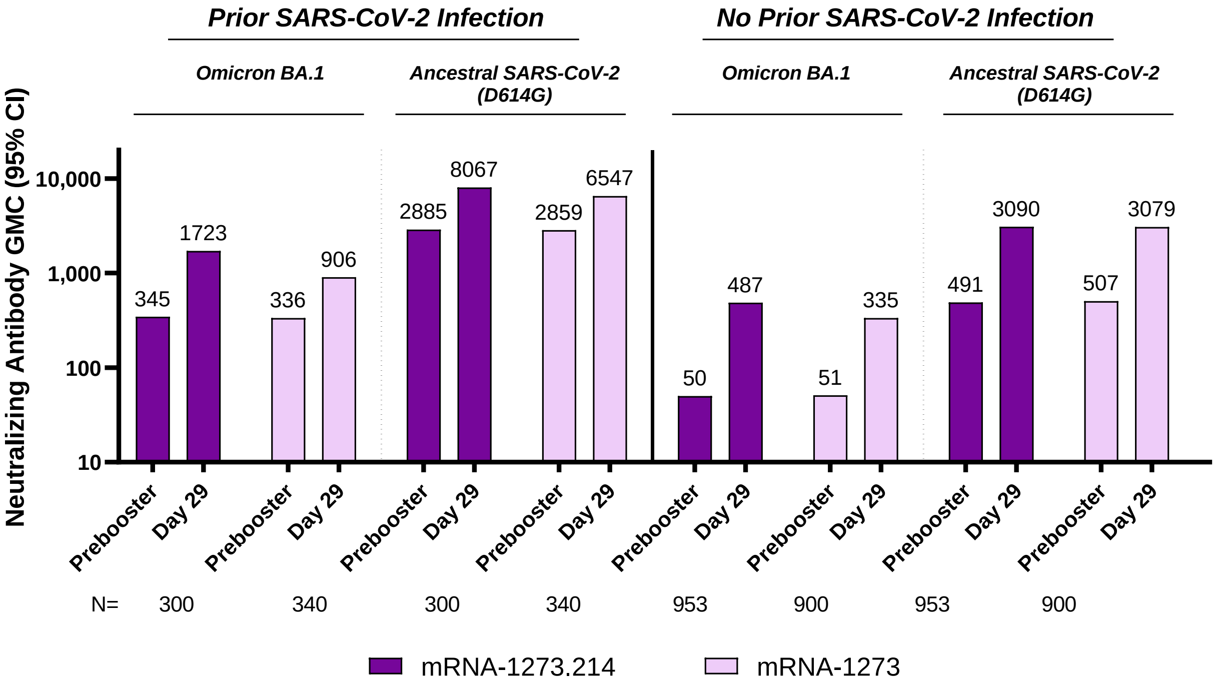


**Figure S6.** The geometric mean concentrations (GMC) of pseudovirus neutralizing antibodies against the omicron BA.1 or ancestral SARS-CoV-2 (D614G) strains were determined in participants in part 2 with or without previous SARS-CoV-2 infection before the booster (per-protocol immunogenicity set). Data are from participants with non-missing data at the time point. Serum samples were collected at day 1 (pre-booster) before the booster dose of 50-µg of mRNA-1273.214 or mRNA-1273 and at 28 days after the booster injection (day 29). N is the number of participants with non-missing data at the timepoint (pre-booster or day 29), CI=confidence interval, GMC=geometric mean concentration, LLOQ=lower limit of quantification, ULOQ=upper limit of quantification. Antibody values assessed by pseudovirus neutralizing antibody assay reported as below the LLOQ (8 for omicron BA.1 and 10 for ancestral SARS-CoV-2 [D614G]) are replaced by 0.5 × LLOQ. Values greater than ULOQ (41,984 for omicron BA.1 and 4,505,600 for ancestral SARS-CoV-2 [D614G]) are replaced by the ULOQ if actual values are not available. 95% CI is calculated based on the t-distribution of the log-transformed values or the difference in the log-transformed values for GM value, then back transformed to the original scale.

**Figure S7. Observed Spike-Binding Antibodies Against Ancestral SARS-CoV-2 and Variants After Receipt of mRNA-1273.529 or mRNA-1273 Boosters, Part 1**

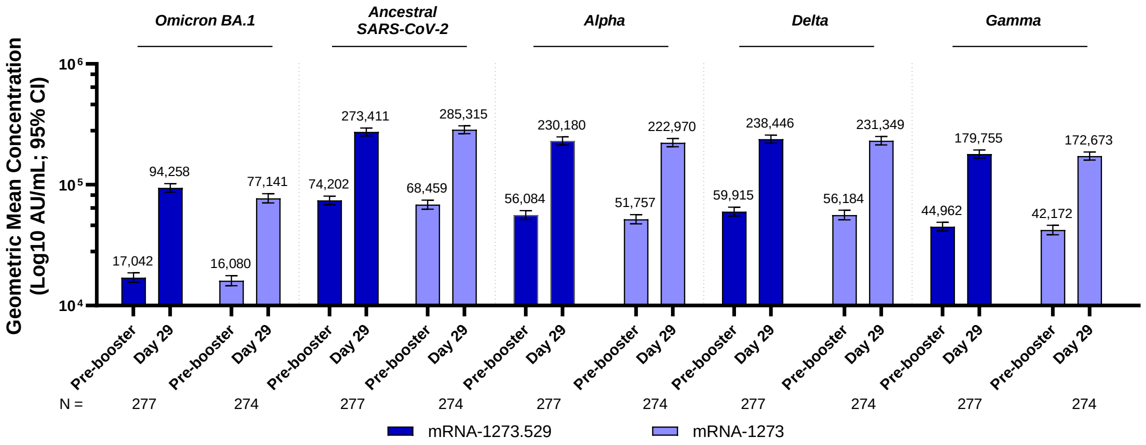


**Figure S7.** The geometric mean concentrations (AU/mL) of antibodies that specifically bind to the omicron BA.1, ancestral SARS-CoV-2, alpha, delta, or gamma spike proteins were determined by the Mesoscale Discovery (MSD) Multiplex (VAC123) assay in participants without previous SARS-CoV-2 infection before the booster (per-protocol immunogenicity – SARS-CoV-2-negative set). Data are from participants in part 2 with non-missing data at the time point. Serum samples were collected at day 1 (pre-booster) before the booster injection dose of 50-µg of mRNA-1273.529 (n=274) or mRNA-1273 (n=277) and at 28 days after the booster injection (day 29). Antibody values reported as below the lower limit of quantification (LLOQ: 102 for omicron BA.1, 79 for ancestral SARS-CoV-2, 52 for alpha, 150 for delta, 143 for gamma) were replaced by 0.5 times the LLOQ. Antibody values reported as greater than the upper limit of quantification (ULLQ:  1,180,000 for omicron BA.1, 5,800,000 for ancestral SARS-CoV-2, 8,800,000 for alpha, 8,000,000 for delta, 5,800,000 for gamma) were converted to the ULOQ if the actual values were not available. Error bars are 95% confidence intervals (CI). The 95% CI were calculated based on the t-distribution of the log-transformed values for the geometric mean value then back transformed to the original scale for presentation.

**Figure S8. Observed Spike-Binding Antibodies Against SARS-CoV-2 Variants and Ancestral SARS-CoV-2 After Receipt of mRNA-1273.214 or mRNA-1273 Administered as Second Boosters, Part 2**

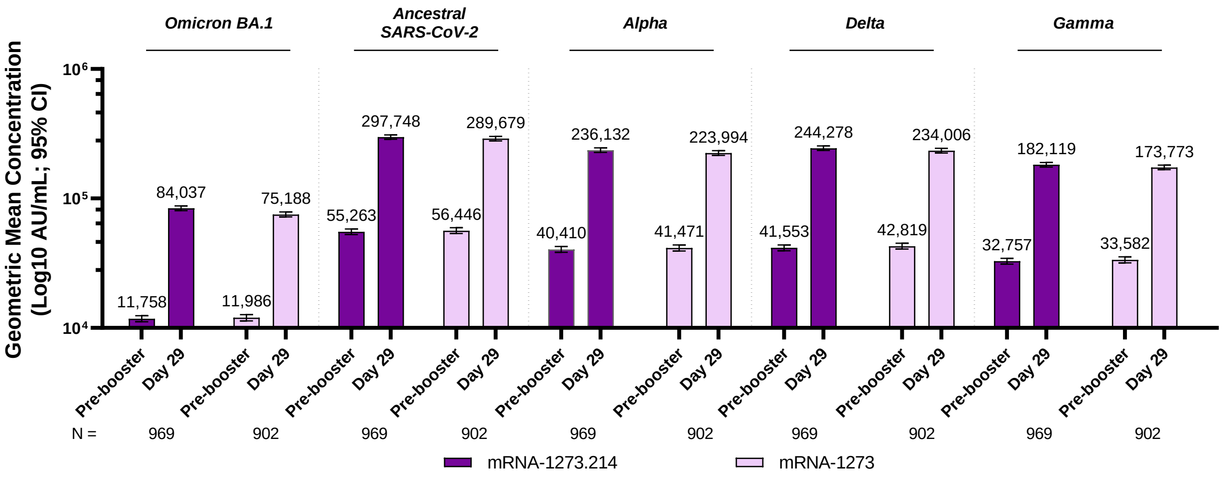


**Figure S8.** The geometric mean concentrations (AU/mL) of antibodies that specifically bind to the omicron BA.1, ancestral SARS-CoV-2, alpha, delta, or gamma spike proteins were determined by the Mesoscale Discovery (MSD) Multiplex (VAC123) assay in participants without previous SARS-CoV-2 infection before the booster (per-protocol immunogenicity – SARS-CoV-2-negative set). Data are from participants in part 2 with non-missing data at the time point. Serum samples were collected at day 1 (pre-booster) before the booster injection dose of 50-µg of mRNA-1273.214 (n=969) or mRNA-1273 (n=902) and at 28 days after the booster injection (day 29). Antibody values reported as below the lower limit of quantification (LLOQ: 102 for omicron BA.1, 79 for ancestral SARS-CoV-2, 52 for alpha, 150 for delta, 143 for gamma) were replaced by 0.5 times the LLOQ. Antibody values reported as greater than the upper limit of quantification (ULLQ: 1,180,000 for omicron BA.1, 5,800,000 for ancestral SARS-CoV-2, 8,800,000 for alpha, 8,000,000 for delta, 5,800,000 for gamma) were converted to the ULOQ if the actual values were not available. Error bars are 95% confidence intervals (CIs). The 95% CIs were calculated based on the t-distribution of the log-transformed values for the geometric mean value then back transformed to the original scale for presentation.

**Figure S9. Omicron Variant Sequences Obtained from Nasopharyngeal Swabs of Positive SARS-CoV-2-infection Cases, Parts 1 and 2**

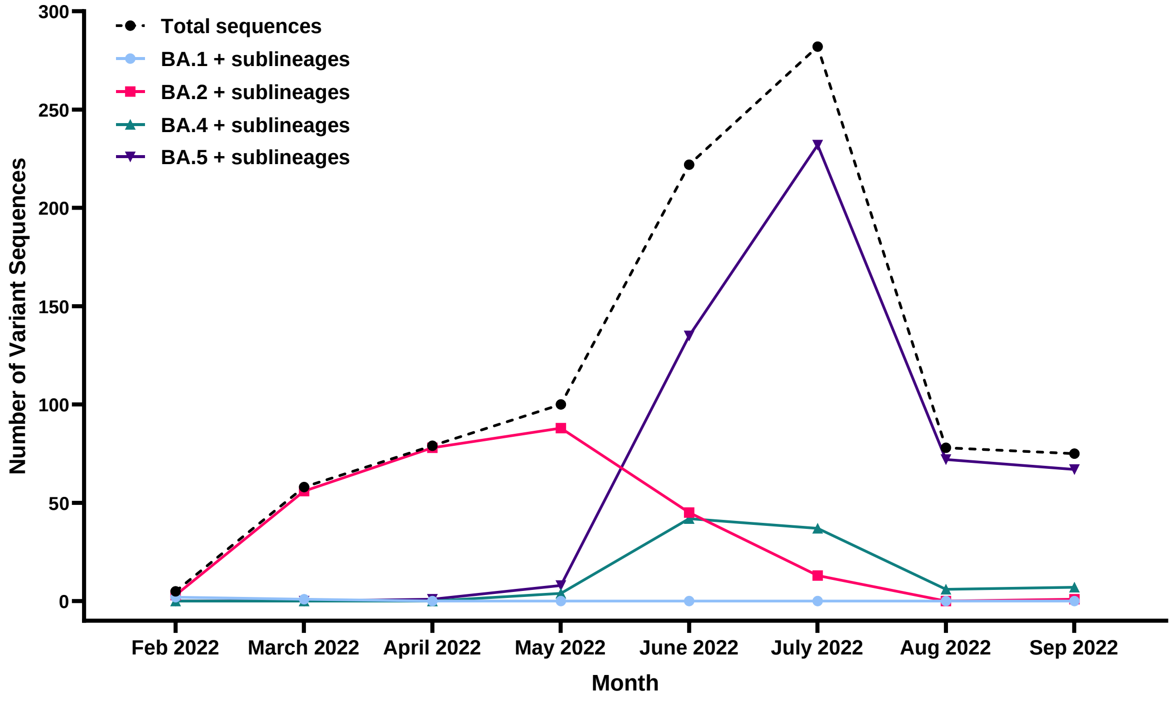


**Figure S9.** Variant sequences (RNA) obtained by RT-PCR in SARS-CoV-2 positive cases with available sequence in parts 1 and 2. The monthly numbers indicate the dates at which the sequences were available and not necessarily when they were collected.

#### Figure S10. Cumulative Event Rates of Covid-19 for BA.2, BA.4 and BA.5 Variants, Part 2

A. BA.2 and sublineages

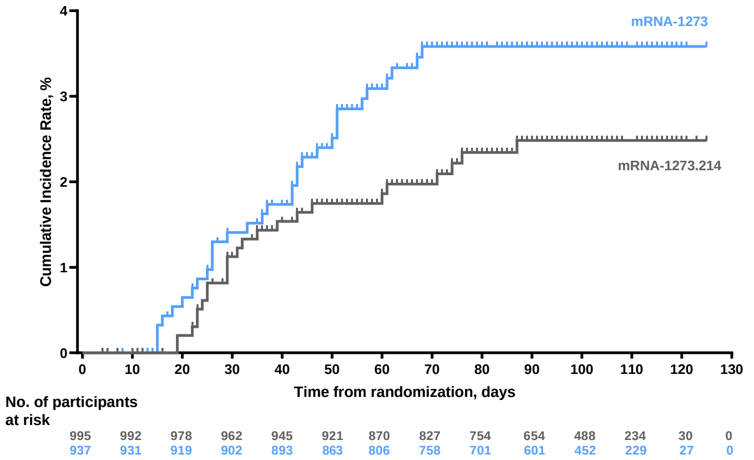


B. BA.4 and sublineages

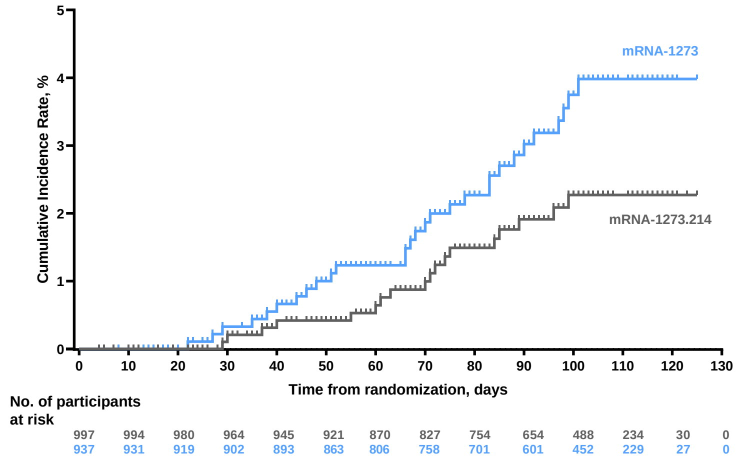


C. BA.5 and sublineages

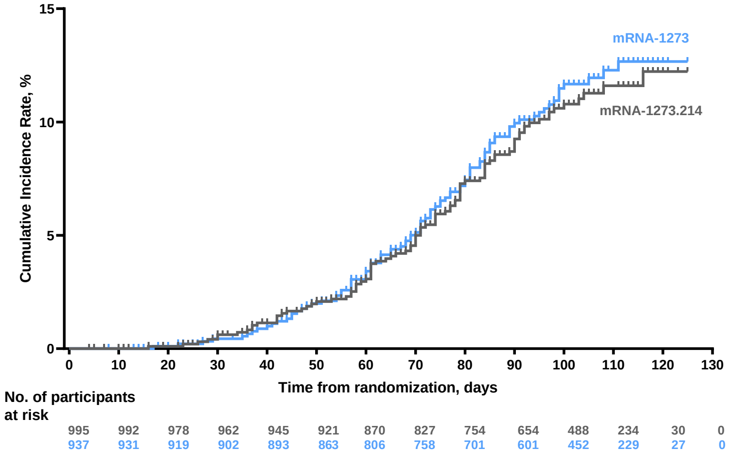

#### Table S1. Objectives and Endpoints
#### Part 1

| Objectives | Endpoints |
|---|---|
| **Primary** | |
| To demonstrate non- inferiority of the immune response of mRNA- 1273.529 compared to mRNA-1273 booster administered as a 4th dose against the omicron BA.1 strain at Day 29 or Month 3 | • Geometric mean concentration (GMC) of mRNA-1273.529 and mRNA-1273 against the BA.1 strain at Day 29 and Month 3 after study vaccine administration<br>• Ratio of $GMC_{mRNA\text{-}1273.529}/GMC_{mRNA\text{-}1273}$ against the BA.1 strain at Day 29 and Month 3 after study vaccine administration |
| To evaluate the safety and reactogenicity of mRNA- 1273.529 and mRNA-1273 administered as a booster dose | • Solicited local and systemic reactogenicity ARs during a 7-day follow up period after vaccination<br>• Unsolicited AEs during the 28-day follow-up period after vaccination<br>• SAEs, medically attended AEs (MAAEs), AEs leading to withdrawal, and AEs of special interest (AESIs) from Day 1 to end of study |
| **Secondary** | |
| **Key Secondary** | |
| To demonstrate superiority of the immune response of mRNA-1273.529 compared to mRNA-1273 administered as a 4th dose against the omicron BA.1 strain at Day 29 or Month 3 | • GMC of mRNA-1273.529 and mRNA-1273 against the BA.1 strain at Day 29 and Month 3 after study vaccine administration<br>• Ratio of $GMC_{mRNA\text{-}1273.529}/GMC_{mRNA\text{-}1273}$ against the BA.1 strain at Day 29 and Month 3 after study vaccine administration |
| **Other Secondary** | |
| To demonstrate non- inferiority of the immune response of mRNA- 1273.529 compared to mRNA-1273 booster administered as a 4th dose against both the omicron BA.1 and the prototype strain at all evaluable time points | • GMC of mRNA-1273.529 and mRNA-1273 against both the BA.1 and the prototype strain at all evaluable time points after study vaccine administration<br>• Ratio of $GMC_{mRNA\text{-}1273.529}/GMC_{mRNA\text{-}1273}$ against both the BA.1 and the prototype strain at all evaluable time points after study vaccine administration |
| To evaluate the seroresponse rate (SRR) of mRNA-1273.529 and mRNA-1273 boosters administered as a 4th dose | • SRR against the BA.1 strain<br>• SRR against the prototype strain |
| **Exploratory Objectives** | |
| To assess for symptomatic and asymptomatic SARS-CoV-2 infection after vaccination with mRNA-1273.529 booster or mRNA-1273 booster | Reverse transcriptase polymerase-chain reaction (RT-PCR)-confirmed symptomatic or asymptomatic SARS-CoV-2 infection will be defined in participants based on the following criteria:<br>• Protocol-defined Covid-19 case definition<br>○ The participant must have experienced at least TWO of the following systemic symptoms: Fever (≥ 38ºC), chills, myalgia, headache, sore throat, new olfactory and taste disorder(s), OR<br>○ The participant must have experienced at least ONE of the following respiratory signs/symptoms: cough, shortness of breath or difficulty breathing, OR clinical or radiographical evidence of pneumonia; AND<br>○ The participant must have at least one nasopharyngeal (NP) swab, nasal swab, or saliva sample (or respiratory sample, if hospitalized) positive for SARS-CoV-2 by RT-PCR.<br>• Center for Disease Control (CDC)-defined Covid-19 definition based on the CDC criteria: the presence of one of the CDC-listed symptoms (https://www.cdc.gov/coronavirus/2019-ncov/symptoms- testing/symptoms.html) and a positive RT-PCR test on a respiratory sample<br>• Asymptomatic SARS-CoV-2 infection is defined as a positive RT-PCR test on a respiratory sample in the absence of symptoms or a positive serologic test for |

| | |
|---|---|
| | antinucleocapsid antibody after a negative test at time of enrollment |
| To evaluate the immunogenicity of mRNA-1273.529 booster against other variant strains | • GMC of mRNA-1273.529 against other variant strains (eg, Alpha, Beta, Delta) after study vaccine administration<br>• Ratio of $GMC_{mRNA-1273.529}/GMC_{mRNA-1273}$ against other variant strains after study vaccine administration<br>• GMFR of mRNA-1273.529 against other variant strains after study vaccine administration SRR against other variant strains |
| To evaluate the genetic and/or phenotypic relationships of isolated SARS-CoV-2 strains to the vaccine sequence | • Characterize the SARS-CoV-2 spike genetic sequence of viral isolates and compare with the vaccine sequence<br>• Characterize the immune responses to vaccine breakthrough isolates |
| To evaluate the genetic and/or phenotypic relationships of isolated SARS-CoV-2 strains to the vaccine sequence | • Characterize the SARS-CoV-2 spike genetic sequence of viral isolates and compare with the vaccine sequence<br>• Characterize the immune responses to vaccine breakthrough isolates |
| Abbreviations: AE = adverse event; AESI = adverse event of special interest; AR = adverse reaction; CDC = Centers for Disease Control and Prevention; GMFR = geometric mean fold rise; GMR = geometric mean concentration ratio; GMC = geometric mean concentration; MAAE = medically attended adverse event; mRNA = messenger ribonucleic acid; NP = nasopharyngeal; RT-PCR = reverse transcription polymerase chain reaction; SAE = serious adverse event; SARS- CoV-2 = severe acute respiratory syndrome coronavirus 2; SRR = seroresponse rate. | |

#### Part 2

| Objectives | Endpoints |
|---|---|
| **Primary** | |
| To demonstrate non-inferiority of the immune response of mRNA- 1273.214 compared to mRNA-1273 booster administered as a 4th dose against the BA.1 strain at Day 29 or Month 3 | • Geometric mean concentration (GMC) of mRNA-1273.214 and mRNA-1273 against the BA.1 strain at Day 29 or Month 3 after study vaccine administration<br>• Ratio of $GMC_{mRNA-1273.214}/GMC_{mRNA-1273}$ against the BA.1 strain at Day 29 or Month 3 after study vaccine administration |
| To demonstrate non-inferiority of the immune response of mRNA-1273.214 compared to mRNA-1273 booster administered as a 4th dose against the prototype strain at Day 29 or Month 3 | • Geometric mean concentration (GMC) of mRNA-1273.214 and mRNA-1273 against the prototype strain at Day 29 or Month 3 after study vaccine administration<br>• Ratio of $GMC_{mRNA-1273.214}/GMC_{mRNA-1273}$ against the prototype strain at Day 29 or Month 3 after study vaccine administration |
| To demonstrate superiority of the immune response of mRNA- 1273.214 compared to mRNA-1273 booster administered as a 4th dose against the BA.1 strain at Day 29 or Month 3 | • GMC of mRNA-1273.529 and mRNA-1273 against the BA.1 strain at Day 29 or Month 3 after study vaccine administration<br>• Ratio of $GMC_{mRNA-1273.529}/GMC_{mRNA-1273}$ against the BA.1 strain at Day 29 or Month 3 after study vaccine administration |
| To evaluate the safety and reactogenicity of mRNA-1273.214 and mRNA-1273 administered as a booster dose | • Solicited local and systemic reactogenicity ARs during a 7-day follow up period after vaccination<br>• Unsolicited AEs during the 28-day follow-up period after vaccination<br>• SAEs, medically attended AEs (MAAEs), AEs leading to withdrawal, and AEs of special interest (AESIs) from Day 1 to end of study |
| **Secondary** | |
| To evaluate the seroresponse rate (SRR) of mRNA-1273.214 and mRNA-1273 boosters administered as a 4th dose | • SRR against the BA.1 strain<br>• SRR against the prototype strain |
| To evaluate the immune response of mRNA-1273.214 compared to mRNA-1273 booster administered as a 4th dose against other variant strains at Day 29 or Month 3 | • GMC of mRNA-1273.214 and mRNA-1273 against other variant strains at Day 29 or Month 3<br>• Ratio of $GMC_{mRNA-1273.214}/GMC_{mRNA-1273}$ against other variant strains at Day 29 or Month 3<br>• SRR against other variant strains |

| | |
|---|---|
| To assess for symptomatic and asymptomatic SARS-CoV-2 infection after vaccination with mRNA-1273.214 booster or mRNA-1273 booster | Reverse transcriptase polymerase-chain reaction (RT-PCR)-confirmed symptomatic or asymptomatic SARS-CoV-2 infection will be defined in participants based on the following criteria:<br>• Protocol-defined Covid-19 case definition<br>○ The participant must have experienced at least TWO of the following systemic symptoms: Fever (≥ 38ºC), chills, myalgia, headache, sore throat, new olfactory and taste disorder(s), OR<br>○ The participant must have experienced at least ONE of the following respiratory signs/symptoms: cough, shortness of breath or difficulty breathing, OR clinical or radiographical evidence of pneumonia; AND<br>○ The participant must have at least one nasopharyngeal (NP) swab, nasal swab, or saliva sample (or respiratory sample, if hospitalized) positive for SARS-CoV-2 by RT-PCR.<br>• Center for Disease Control (CDC) defined Covid-19 definition based on the CDC criteria: the presence of one of the CDC listed symptoms (https://www.cdc.gov/coronavirus/2019-ncov/symptoms-testing/symptoms.html) and a positive RT-PCR test on a respiratory sample<br>• Asymptomatic SARS-CoV-2 infection is defined as a positive RT-PCR test on a respiratory sample in the absence of symptoms or a positive serologic test for antinucleocapsid antibody after a negative test at time of enrollment |
| **Exploratory Objectives** | |
| To evaluate cellular immunogenicity in a subset of participants | • Frequency, magnitude, and phenotype of virus-specific T-cell and B-cell responses measured by flow cytometry or other methods, and to perform targeted repertoire analysis of T-cells and B-cells after vaccination |
| To evaluate the genetic and/or phenotypic relationships of isolated SARS-CoV-2 strains to the vaccine sequence | • Characterize the SARS-CoV-2 spike genetic sequence of viral isolates and compare with the vaccine sequence |
| To evaluate the immunogenicity of mRNA-1273.214 booster compared to mRNA-1273 booster administered as a 4th dose study vaccine in participants at Month 6 | • GMC of mRNA-1273.214 against both the BA.1 and the prototype strain at Month 6 after study vaccine administration<br>• Ratio of GMCmRNA-1273.214/GMCmRNA-1273 against the BA.1 strain at Month 6 after study vaccine administration<br>• Ratio of $GMC_{mRNA\text{-}1273.214}/GMC_{mRNA\text{-}1273}$ against the prototype strain at Month 6 after study vaccine administration<br>• GMFR of mRNA-1273.214 and mRNA-1273 against the BA.1 and prototype strain at all evaluable timepoints after study vaccine administration<br>• SRR against both the BA.1 and prototype strain at Month 6 |
| Abbreviations: AE = adverse event; AESI = adverse event of special interest; AR = adverse reaction; CDC = Centers for Disease Control and Prevention; GMFR = geometric mean fold rise; GMR = geometric mean concentration ratio; GMC = geometric mean concentration; MAAE = medically attended adverse event; mRNA = messenger ribonucleic acid; NP = nasopharyngeal; RT-PCR = reverse transcription polymerase chain reaction; SAE = serious adverse event; SARS- CoV-2 = severe acute respiratory syndrome coronavirus 2; SRR = seroresponse rate. | |

#### Table S2. Analysis Populations

| Set | Description |
| --- | --- |
| Full Analysis Set (FAS) | The FAS consists of all randomized participants who received the study vaccination. Participants are analyzed according to their randomized study arm. |
| Modified Intent-to-Treat (mITT) Set | The mITT Set consists of all participants in the FAS who have no serologic or virologic evidence of prior SARS-CoV-2 infection (both negative RT-PCR test for SARS-CoV-2 and negative serology test based on bAb specific to SARS-CoV-2 nucleocapsid) prebooster, i.e., all FAS participants with pre- booster/baseline SARS-CoV-2 negative status. Participants are analyzed according to their randomized study arm. |
| PP Set for Immunogenicity (PPSI) | The PPSI consists of all participants in the FAS who received the planned dose of study vaccination and have no major protocol deviations that impact key or critical data. Participants are analyzed according to their randomized study arm. |
| PP Set for Immunogenicity – SARS-CoV-2 negative (PPSI-Neg) | Participants in the PPSI who have no serologic or virologic evidence of SARS-CoV-2 infection up to day of visit analysis, i.e., who are SARS-CoV-2 negative, defined by both negative RT- PCR test for SARS-CoV-2 and negative serology test based on bAb specific to SARS-CoV-2 nucleocapsid. For this interim analysis, the PPSI-Neg required no serologic or virologic evidence of SARS-CoV-2 infection up to and including the Day 29 assessment.<br><br>PPSI-Neg is the primary analysis set for analyses of immunogenicity unless otherwise specified. |
| Solicited Safety Set | The Solicited Safety Set consists of all randomized participants who received study vaccine and contribute any solicited AR data.<br><br>The Solicited Safety Set is used for the analyses of solicited ARs. Participants will are included in the study arm that they actually received. |
| Safety Set | The Safety Set consists of all randomized participants who received study vaccine. The Safety Set is used for all analyses of safety except for the solicited ARs. Participants areincluded in the study arm that they actually received. |
| PP Set for Efficacy (PPSE) | The PPSE consists of all participants in the mITT who received the planned study vaccination and had no major protocol deviations that impact key or critical data. |

**Table S3. Observed Neutralizing Antibody Geometric Mean Concentrations Against Ancestral SARS-CoV-2 (D614G) and Omicron BA.1 after Receipt of mRNA-1273.214 or mRNA-1273 Administered as Second Boosters by Prior SARS-CoV-2 Infection Status at Pre-Booster, Part 2**

| | All participants* | | No prior SARS-CoV-2 infection | | Prior SARS-CoV-2 infection | |
|---|---|---|---|---|---|---|
| | mRNA-1273.214 50 µg | mRNA-1273 50 µg | mRNA-1273.214 50 µg | mRNA-1273 50 µg | mRNA-1273.214 50 µg | mRNA-1273 50 µg |
| **Omicron BA.1** | **N=1334** | **N=1311** | **N=953** | **N=900** | **N=300** | **N=340** |
| Pre-booster, n | 1330 | 1310 | 949 | 899 | 300 | 340 |
| Observed GMC (95% CI) | 77.7 (72.3-83.5) | 84.0 (78.1-90.5) | 50.4 (47.2-53.7) | 51.2 (47.8-54.9) | 345.4 (299.1-399.0) | 335.8 (295.3-381.8) |
| Day 29, n | 1327 | 1299 | 949 | 893 | 297 | 336 |
| Observed GMC (95% CI) | 644.2 (605.9-685.0) | 431.6 (407.9-456.7) | 486.9 (456.0-519.8) | 335.4 (315.1-357.0) | 1722.5 (1549.2-1915.2) | 905.8 (820.9-999.6) |
| GMFR (95% CI) | 8.3 (7.9-8.8) | 5.1 (4.9-5.4) | 9.7 (9.2-10.2) | 6.5 (6.1-6.9) | 5.0 (4.5-5.5) | 2.7 (2.5-2.9) |
| **Ancestral SARS-CoV-2 (D614G)** | **N=1334** | **N=1311** | **N=953** | **N=900** | **N=300** | **N=340** |
| Pre-booster, n | 1315 | 1283 | 942 | 888 | 298 | 330 |
| Observed GMC (95% CI) | 735.8 (688.4-786.4) | 796.4 (745.2-851.2) | 491.3 (462.1-522.4) | 507.2 (476.8-539.7) | 2884.7 (2561.3-3248.9) | 2858.6 (2573.1-3175.8) |
| Day 29, n | 1308 | 1285 | 940 | 887 | 289 | 330 |
| Observed GMC (95% CI) | 3811.1 (3609.7-4023.6) | 3716.6 (3532.0-3910.9) | 3090.1 (2907.1-3284.6) | 3078.8 (2905.2-3262.6) | 8067.2 (7367.9-8832.8) | 6546.9 (5988.3-7157.6) |
| GMFR (95% CI) | 5.3 (5.0-5.6) | 4.7 (4.5-5.0) | 6.3 (5.9-6.7) | 6.0 (5.7-6.4) | 2.9 (2.6-3.1) | 2.3 (2.1-2.6) |

n = number of participants with non-missing data at the timepoint (pre-booster or Day 29), CI=confidence interval, GMC=geometric mean concentration, GMFR=geometric mean fold rise (day 29 compared with pre-booster), LLOQ=lower limit of quantification, ULOQ=upper limit of quantification. Antibody values assessed by pseudovirus neutralizing antibody assay reported as below the LLOQ (8 for omicron BA.1 and 10 for ancestral SARS-CoV-2 [D614G]) are replaced by 0.5 × LLOQ. Values greater than ULOQ (41,984 for omicron BA.1 and 4,505,600 for ancestral SARS-CoV-2 [D614G] or) are replaced by the ULOQ if actual values are not available. 95% CI is calculated based on the t-distribution of the log-transformed values or the difference in the log-transformed values for GMC and GMFR, respectively, then back transformed to the original scale.
*All participants include those with missing SARS-CoV-2 status at pre-booster (n=81 for mRNA-1273.214 group; n=71 for mRNA-1273 group).

**Table S4. Observed Binding Antibody Geometric Mean Concentrations and Geometric Mean Ratios Against Ancestral SARS-CoV-2 and Variants after Receipt of mRNA-1273.529 or mRNA-1273 Administered as Second Boosters in Participants with No Evidence of SARS-CoV-2 Infection at Pre-Booster, Part 1**

#### A. Ancestral SARS-CoV-2 and Omicron BA.1

| | Omicron BA.1 | | Ancestral SARS-CoV-2 | |
|---|---|---|---|---|
| | **mRNA-1273.529 50 µg N=274** | **mRNA-1273 50 µg N=277** | **mRNA-1273.529 50 µg N=274** | **mRNA-1273 50 µg N=277** |
| Pre-booster, n† | 273 | 277 | 273 | 277 |
| Observed GMC (95% CI)§ | 17,042 (15,551-18,675) | 16,080 (14,606-17,704) | 74,202 (68,281-80,637) | 68,459 (62,755-74,681) |
| Day 29, n† | 274 | 277 | 274 | 277 |
| Observed GMC (95% CI)§ | 94,258 (86,921-102,215) | 77,141 (70,723-84,141) | 273,411 (253,644-294,718) | 285,315 (264,669-307,572) |
| GMFR (95% CI)§ | 5.5 (5.2-5.9) | 4.8 (4.5-5.2) | 3.7 (3.5-3.9) | 4.2 (3.9-4.5) |
| Estimated GMC (95% CI)¶ | 92,367 (85,043-100,321) | 78,428 (72,267-85,113) | 265,905 (251,330-281,324) | 291,613 (275,779-308,356) |
| GMR (95% CI)¶ | 1.18 (1.05-1.32) | | 0.91 0.84-0.99) | |

#### B. Other Variants

| | Alpha | | Delta | | Gamma | |
|---|---|---|---|---|---|---|
| | **mRNA-1273.529 50 µg N=274** | **mRNA-1273 50 µg N=277** | **mRNA-1273.529 50 µg N=274** | **mRNA-1273 50 µg N=277** | **mRNA-1273.529 50 µg N=274** | **mRNA-1273 50 µg N=277** |
| Pre-booster, n† | 273 | 277 | 273 | 277 | 273 | 277 |
| Observed GMC (95% CI)§ | 56,084 (51,540-61,028) | 51,757 (47,359-56,563) | 59,915 (55,087-65,167) | 56,184 (51,225-61,622) | 44,962 (41,411-48,817) | 42,172 (38,543-46,141) |
| Day 29, n† | 274 | 277 | 274 | 277 | 274 | 277 |
| Observed GMC (95% CI)§ | 230,180 (213,176-248,540) | 222,970 (206,101-220,485) | 238,446 (220,938-257,343) | 231,349 (213,697-250,459) | 179,755 (166,532-194,028) | 172,673 (159,904-186,462) |
| GMFR (95% CI)§ | 4.1 (3.9-4.4) | 4.3 (4.0-4.6) | 4.0 (3.7-4.2) | 4.1 (3.8-4.4) | 4.0 (3.7-4.3) | 4.1 (3.8-4.4) |
| Estimated GMC (95% CI)¶ | 223,601 (211,027-236,924) | 227,871 (215,175-241,317) | 232,758 (219,873-246,398) | 235,143 (222,248-248,786) | 175,390 (165,639-185,715) | 175,424 (165,764-185,647) |
| GMR (95% CI)¶ | 0.98 (0.91-1.06) | | 0.99 (0.92-1.07) | | 1.00 (0.92-1.08) | |

CI = confidence interval; GMC = geometric mean concentration; GMFR = geometric mean fold -rise, GMR = geometric mean ratio (mRNA-1273.529 relative to mRNA-1273); LLOQ, lower limit of quantification; MSD = Meso Scale Discovery; ULOQ = upper limit of quantification. Binding antibody values assessed by MSD assay reported as below the LLOQ (69 for ancestral SARS-CoV-2; 102 for omicron BA.1; 52 for alpha; 150 for delta; 143 for gamma) were replaced by 0.5 × LLOQ. Values greater than ULOQ (1,180,000 for omicron BA.1; 14,400,000 for ancestral SARS-CoV-2; 8,800,000 for alpha; 8,000,000 for delta; 5,800,000 for gamma;) were replaced by the ULOQ if actual values are not available.
†Number of participants with non-missing data at the timepoint (baseline or post-baseline).
§95% CI based on the t-distribution of log-transformed values or difference in the log-transformed values for GMC value and GMFR, respectively, then back transformed to the original scale.
¶ The log-transformed antibody levels are analyzed using an analysis of covariance (ANCOVA) model with the treatment variable as fixed effect, adjusting for age group (≥16 to <65, ≥65 years), most recent Covid-19 vaccination type (mRNA, viral vector), pre-booster antibody level, and pre-booster SARS-CoV-2 status (negative, positive, or missing). The LS means, difference of LS means, and 95% CI are back transformed to the original scale for presentation.

**Table S5. Observed Binding Antibody Geometric Mean Concentrations and Geometric Mean Ratios Against Ancestral SARS-CoV-2 and Variants after Receipt of mRNA-1273.214 or mRNA-1273 Administered as Second Boosters in Participants with No Evidence of SARS-CoV-2 Infection at Pre-Booster, Part 2**

#### A. Ancestral SARS-CoV-2 and Omicron BA.1

| | Omicron BA.1 | | Ancestral SARS-CoV-2 | |
|---|---|---|---|---|
| | **mRNA-1273.214 50 µg N=969** | **mRNA-1273 50 µg N=902** | **mRNA-1273.214 50 µg N=969** | **mRNA-1273 50 µg N=902** |
| Pre-booster, n† | 969 | 902 | 969 | 902 |
| Observed GMC§ | 11,758 (11,136-12,415) | 11,986 (11,335-12,675) | 55,263 (52,584-58,080) | 56,446 (53,629-59,411) |
| Day 29, n† | 967 | 902 | 967 | 902 |
| Observed GMC (95% CI)§ | 84,037 (80,572-87,651) | 75,188 (71,911-78,615) | 297,748 (286,395-309,551) | 289,679 (278,124-301,715) |
| GMFR (95% CI)§ | 7.1 (6.8-7.5) | 6.3 (6.0-6.6) | 5.4 (5.2-5.6) | 5.1 (4.9-5.4) |
| Estimated GMC (95% CI)¶ | 79,177 (59,121-106,037) | 70,193 (52,406-94,016) | 284,300 (215,947-374,289) | 273,980 (208,085-360,741) |
| GMR (95% CI)¶ | 1.13 (1.06-1.21) | | 1.04 (0.98-1.10) | |

#### B. Other Variants

| | Alpha | | Delta | | Gamma | |
|---|---|---|---|---|---|---|
| | **mRNA-1273.214 50 µg N=969** | **mRNA-1273 50 µg N=902** | **mRNA-1273.214 50 µg N=969** | **mRNA-1273 50 µg N=902** | **mRNA-1273.214 50 µg N=969** | **mRNA-1273 50 µg N=902** |
| Pre-booster, n† | 969 | 902 | 969 | 902 | 969 | 902 |
| Observed GMC§ | 40,410 (38,424-42,499) | 41,471 (39,354-43,702) | 41,553 (39,441-43,778) | 42,819 (40,625-45,132) | 32,757 (31,116-34,485) | 33,582 (31,854-35,403) |
| Day 29, n† | 967 | 902 | 967 | 902 | 967 | 902 |
| Observed GMC (95% CI)§ | 236,132 (226,731-245,923) | 223,994 (214,782-233,601) | 244,278 (234,536-254,425) | 234,006 (224,329-244,099) | 182,119 (174,972-189,557) | 173,773 (166,638-181,213) |
| GMFR (95% CI)§ | 5.8 (5.6-6.1) | 5.4 (5.2-5.7) | 5.9 (5.6-6.1) | 5.5 (5.2-5.7) | 5.6 (5.3-5.8) | 5.2 (4.9-5.4) |
| Estimated GMC (95% CI)¶ | 231,088 (186,771-285,920) | 216,496 (174,963-267,889) | 240,481 (194,771-296,919) | 227,025 (183,859-280,327) | 172,280 (139,548-212,690) | 162,487 (131,603-200,618) |
| GMR (95% CI)¶ | 1.07 (1.02-1.12) | | 1.06 (1.01-1.11) | | 1.06 (1.01-1.11) | |

CI = confidence interval; GMC = geometric mean concentration; GMFR = geometric mean fold -rise, GMR = geometric mean ratio (mRNA-1273.214 relative to mRNA-1273); LLOQ, lower limit of quantification; MSD = Meso Scale Discovery; ULOQ = upper limit of quantification. Binding antibody values assessed by MSD assay reported as below the LLOQ (102 for omicron BA.1; 69 for ancestral SARS-CoV-2; 52 for alpha; 150 for delta; 143 for gamma) were replaced by 0.5 × LLOQ. Values greater than ULOQ (1,180,000 for omicron BA.1;14,400,000 for ancestral SARS-CoV-2; 8,800,000 for alpha; 8,000,000 for delta; 5,800,000 for gamma) were replaced by the ULOQ if actual values are not available.
†Number of participants with non-missing data at the timepoint (baseline or post-baseline).
§95% CI based on the t-distribution of log-transformed values or difference in the log-transformed values for GMC value and GMFR, respectively, then back transformed to the original scale.
¶ The log-transformed antibody levels are analyzed using an analysis of covariance (ANCOVA) model with the treatment variable as fixed effect, adjusting for age group (≥16 to <65, ≥65 years), most recent Covid-19 vaccination type (mRNA, viral vector), pre-booster antibody level, and pre-booster SARS-CoV-2 status (negative, positive, or missing). The LS means, difference of LS means, and 95% CI are back transformed to the original scale for presentation.

#### Table S6. Solicited Local and Systemic Adverse Reactions Within 7 Days After Receipt of of mRNA-1273.214, mRNA-1273.529, or mRNA-1273 Boosters, Solicited Safety Set

| Solicited Adverse Reaction n (%) | Part 1 | | Part 2 | |
|---|---|---|---|---|
| | mRNA 1273.529 50 ug (N=367) | mRNA-1273 50 ug (N=357) | mRNA-1273.214 50 ug (N=1421) | mRNA-1273 50 ug (N=1398) |
| Solicited Adverse Reactions, N1 | 367 | 357 | 1421 | 1398 |
| Any Solicited Adverse Reactions | 335 (91.3) | 333 (93.3) | 1285 (90.4) | 1317 (94.2) |
| Grade 1 | 193 (52.6) | 171 (47.9) | 743 (52.3) | 667 (47.7) |
| Grade 2 | 120 (32.7) | 128 (35.9) | 432 (30.4) | 499 (35.7) |
| Grade 3 | 22 (6.0) | 34 (9.5) | 110 (7.7) | 151 (10.8) |
| Grade 4 | 0 | 0 | 0 | 0 |
| Local Adverse Reactions-N1 | 367 | 357 | 1421 | 1398 |
| Any Local Adverse Reactions | 310 (84.5) | 318 (89.1) | 1187 (83.5) | 1256 (89.8) |
| Grade 1 | 269 (73.3) | 256 (71.7) | 1033 (72.7) | 980 (70.1) |
| Grade 2 | 31 (8.4) | 49 (13.7) | 111 (7.8) | 208 (14.9) |
| Grade 3 | 10 (2.7) | 13 (3.6) | 43 (3.0) | 68 (4.9) |
| Grade 4 | 0 | 0 | 0 | 0 |
| Pain - N1 | 367 | 357 | 1421 | 1398 |
| Any | 307 (83.7) | 315 (88.2) | 1167 (82.1) | 1246 (89.1) |
| Grade 1 | 280 (76.3) | 267 (74.8) | 1071 (75.4) | 1055 (75.5) |
| Grade 2 | 20 (5.4) | 38 (10.6) | 76 (5.3) | 153 (10.9) |
| Grade 3 | 7 (1.9) | 10 (2.8) | 20 (1.4) | 38 (2.7) |
| Grade 4 | 0 | 0 | 0 | 0 |
| Erythema - N1 | 367 | 357 | 1421 | 1398 |
| Any | 15 (4.1) | 13 (3.6) | 58 (4.1) | 102 (7.3) |
| Grade 1 | 6 (1.6) | 8 (2.2) | 37 (2.6) | 41 (2.9) |
| Grade 2 | 7 (1.9) | 5 (1.4) | 14 (1.0) | 48 (3.4) |
| Grade 3 | 2 (0.5) | 0 | 7 (0.5) | 13 (0.9) |
| Grade 4 | 0 | 0 | 0 | 0 |
| Swelling - N1 | 367 | 357 | 1421 | 1398 |
| Any | 29 (7.9) | 30 (8.4) | 110 (7.7) | 169 (12.1) |
| Grade 1 | 17 (4.6) | 13 (3.6) | 64 (4.5) | 93 (6.7) |
| Grade 2 | 8 (2.2) | 15 (4.2) | 30 (2.1) | 50 (3.6) |
| Grade 3 | 4 (1.1) | 2 (0.6) | 16 (1.1) | 26 (1.9) |
| Grade 4 | 0 | 0 | 0 | 0 |
| Axillary Swelling/Tenderness - N1 | 367 | 357 | 1421 | 1398 |
| Any | 63 (17.2) | 60 (16.8) | 248 (17.5) | 252 (18.0) |
| Grade 1 | 55 (15.0) | 52 (14.6) | 218 (15.3) | 212 (15.2) |
| Grade 2 | 8 (2.2) | 5 (1.4) | 26 (1.8) | 36 (2.6) |
| Grade 3 | 0 | 3 (0.8) | 4 (0.3) | 4 (0.3) |
| Grade 4 | 0 | 0 | 0 | 0 |
| Systemic Adverse Reactions - N1 | 367 | 357 | 1421 | 1398 |
| Any Systemic Adverse Reactions | 256 (69.8) | 265 (74.2) | 997 (70.2) | 1052 (75.3) |
| Grade 1 | 128 (34.9) | 118 (33.1) | 510 (35.9) | 476 (34.0) |
| Grade 2 | 115 (31.3) | 123 (34.5) | 411 (28.9) | 468 (33.5) |
| Grade 3 | 13 (3.5) | 24 (6.7) | 76 (5.3) | 108 (7.7) |
| Grade 4 | 0 | 0 | 0 | 0 |
| Fever - N1 | 367 | 357 | 1417 | 1392 |
| Any | 9 (2.5) | 11 (3.1) | 35 (2.5) | 64 (4.6) |
| Grade 1 | 4 (1.1) | 7 (2.0) | 20 (1.4) | 46 (3.3) |
| Grade 2 | 4 (1.1) | 2 (0.6) | 12 (0.8) | 11 (0.8) |
| Grade 3 | 1 (0.3) | 2 (0.6) | 3 (0.2) | 7 (0.5) |
| Grade 4 | 0 | 0 | 0 | 0 |
| Headache - N1 | 367 | 357 | 1421 | 1398 |

| | | | | |
|---|---|---|---|---|
| Any | 167 (45.5) | 175 (49.0) | 649 (45.7) | 700 (50.1) |
| Grade 1 | 129 (35.1) | 120 (33.6) | 496 (34.9) | 502 (35.9) |
| Grade 2 | 36 (9.8) | 49 (13.7) | 131 (9.2) | 172 (12.3) |
| Grade 3 | 2 (0.5) | 6 (1.7) | 22 (1.5) | 26 (1.9) |
| Grade 4 | 0 | 0 | 0 | 0 |
| Fatigue - N1 | 367 | 357 | 1421 | 1398 |
| Any | 202 (55.0) | 215 (60.2) | 810 (57.0) | 862 (61.7) |
| Grade 1 | 103 (28.1) | 93 (26.1) | 416 (29.3) | 384 (27.5) |
| Grade 2 | 91 (24.8) | 108 (30.3) | 346 (24.3) | 395 (28.3) |
| Grade 3 | 8 (2.2) | 14 (3.9) | 48 (3.4) | 83 (5.9) |
| Grade 4 | 0 | 0 | 0 | 0 |
| Myalgia - N1 | 367 | 357 | 1421 | 1398 |
| Any | 147 (40.1) | 153 (42.9) | 538 (37.9) | 596 (42.6) |
| Grade 1 | 82 (22.3) | 83 (23.2) | 309 (21.7) | 299 (21.4) |
| Grade 2 | 62 (16.9) | 62 (17.4) | 203 (14.3) | 255 (18.2) |
| Grade 3 | 3 (0.8) | 8 (2.2) | 26 (1.8) | 42 (3.0) |
| Grade 4 | 0 | 0 | 0 | 0 |
| Arthralgia - N1 | 367 | 357 | 1421 | 1398 |
| Any | 105 (28.6) | 114 (31.9) | 396 (27.9) | 453 (32.4) |
| Grade 1 | 61 (16.6) | 73 (20.4) | 234 (16.5) | 258 (18.5) |
| Grade 2 | 42 (11.4) | 37 (10.4) | 143 (10.1) | 167 (11.9) |
| Grade 3 | 2 (0.5) | 4 (1.1) | 19 (1.3) | 28 (2.0) |
| Grade 4 | 0 | 0 | 0 | 0 |
| Nausea/Vomiting - N1 | 367 | 357 | 1421 | 1398 |
| Any | 40 (10.9) | 42 (11.8) | 152 (10.7) | 178 (12.7) |
| Grade 1 | 36 (9.8) | 34 (9.5) | 129 (9.1) | 140 (10.0) |
| Grade 2 | 4 (1.1) | 8 (2.2) | 23 (1.6) | 38 (2.7) |
| Grade 3 | 0 | 0 | 0 | 0 |
| Grade 4 | 0 | 0 | 0 | 0 |
| Chills - N1 | 367 | 357 | 1421 | 1398 |
| Any | 102 (27.8) | 87 (24.4) | 337 (23.7) | 387 (27.7) |
| Grade 1 | 59 (16.1) | 46 (12.9) | 235 (16.5) | 205 (14.7) |
| Grade 2 | 43 (11.7) | 41 (11.5) | 98 (6.9) | 172 (12.3) |
| Grade 3 | 0 | 0 | 4 (0.3) | 10 (0.7) |
| Grade 4 | 0 | 0 | 0 | 0 |
| Percentages based on the solicited safety set. N1=Number of exposed participants with any information about the adverse event. | | | | |

**Table S7. Summary of Unsolicited Adverse Events within 28 Days after Receipt of the mRNA-1273.214, mRNA-1273.529, or mRNA-1273 Boosters, Safety Set**

| n (%) | Part 1 | | Part 2 | |
|---|---|---|---|---|
| | mRNA-1273.529 50 µg N=367 | mRNA-1273 50 µg N=357 | mRNA-1273.214 50 µg N=1422 | mRNA-1273 50 µg N=1402 |
| ***AEs Regardless of Relationship to Study Vaccination*** | | | | |
| All | 144 (39.2) | 127 (35.6) | 448 (31.5) | 417 (29.7) |
| Serious | 5 (1.4) | 1 (0.3) | 6 (0.4) | 5 (0.4) |
| Fatal | 0 | 0 | 0 | 0 |
| Medically Attended | 65 (17.78) | 60 (16.8) | 166 (11.7) | 142 (10.1) |
| Leading to Study Discontinuation | 1 (0.3) | 0 | 2 (0.1) | 1 (0.1) |
| Grade 3 or Higher | 8 (2.2) | 7 (2.0) | 12 (0.8) | 11 (0.8) |
| ***AEs Related to Study Vaccination*** | | | | |
| All | 27 (7.4) | 30 (8.4) | 69 (4.9) | 71 (5.1) |
| Serious | 2 (0.5)[*] | 0 | 0 | 1 (0.1)[†] |
| Fatal | 0 | 0 | 0 | 0 |
| Medically Attended | 5 (1.4) | 2 (0.6) | 6 (0.4) | 7 (0.5) |
| Leading to Study Discontinuation | 1 (0.3) | 0 | 0 | 0 |
| Grade 3 or Higher | 4 (1.1) | 2 (0.6) | 2 (0.1) | 3 (0.2) |

Numbers are based on actual treatment group and percentages are based on the number of participants in the Safety Set.

*Two (0.5%) SAEs considered related to study vaccination occurred in Part 1 in two participants (60-70 years of age) in the mRNA-1273.529 arm. One participant with a medical history/risk factors including hypertension and asthma experienced a bilateral pulmonary emboli [PE] 14 days after receiving the study vaccine and discontinued from the study on day 82 post-booster. The Investigator considered the event related to the study vaccination given the temporal association and the Sponsor agreed that this could be possibly related given the temporal association; however, to date, there is no known mechanism of action, and evaluation of post-authorization/post-marketing has not demonstrated an increased risk for PE among recipients of mRNA-1273 vaccine. The other participant who had 2 runs of ventricular tachycardia (17 days and 30 days after the study vaccine) had a history of VT leading to cardiac arrests, myocardial infarctions, status postcoronary artery bypass surgery, and implantable (pacemaker with defibrillator) cardiac device.

†One (0.1%) SAE of multiple pulmonary emboli was reported to have occurred within 24 hours of study vaccination in an individual (50-55 years) with a history of deep vein thrombosis (DVT) within 24 hours of the study vaccination and was considered related to study vaccination in the mRNA-1273 control group by the Investigator; however, the Sponsor did not consider this case to be related to vaccination given the short time to onset (within 24 hours of vaccination) and the recent history of DVT, which is a strong confounder/alternate etiology.

#### Table S8. Incidence Rates Starting 14 days after Randomization

| | Part 1 | | Part 2 | |
|---|---|---|---|---|
| | **mRNA-1273.529 50 µg (N=312)** | **mRNA-1273 50 µg (N=312)** | **mRNA-1273.214 50 µg (N=997)** | **mRNA-1273 50 µg (N=937)** |
| Covid-19 cases (COVE definition), n (%) | 76 (24.4) | 86 (27.6) | 158 (15.8) | 166 (17.7) |
| Person years* | 113.3 | 111.8 | 249.6 | 233.3 |
| Incidence rate/1000 person-yrs (95% CI)† | 670.5 (528.3-839.3) | 769.3 (615.4-950.1) | 633.0 (538.1-739.7) | 711.6 (607.5-828.5) |
| Covid-19 cases (CDC-defined), n (%) | 81 (26.0) | 97 (31.1) | 182 (18.3) | 175 (18.7) |
| Person years* | 110.8 | 109.2 | 246.2 | 231.7 |
| Incidence rate/1000 person-yrs (95% CI)† | 731.1 (580.6-908.7) | 888.3 (720.4-1083.7) | 739.2 (635.7-854.8) | 755.4 (647.6-876.0) |
| SARS-CoV-2 infection, n (%) | 93 (29.8) | 107 (34.3) | 243 (24.4) | 248 (26.5) |
| Person years* | 107.8 | 105.6 | 240.5 | 225.6 |
| Incidence rate/1000 person-yrs (95% CI)† | 862.5 (696.2-1056.7) | 1012.9 (830.1-1224.0) | 1010.5 (887.5-1145.9) | 1099.1 (966.5-1244.7) |
| Asymptomatic infection, n (%) | 12 (3.8) | 11 (3.5) | 62 (6.2) | 73 (7.8) |
| Person years* | 108.2 | 105.9 | 240.6 | 225.8 |
| Incidence rate/1000 person-yrs (95% CI)† | 110.9 (57.3-193.8) | 103.9 (51.9-185.9) | 257.7 (197.6-330.4) | 323.3 (253.4-406.5) |

Starting 14 days after randomization.
Covid-19 (COVE definition) protocol-defined case primary case definition, defined as meeting clinical criteria based on both symptoms of Covid-19 and positive RT-PCR test results.[4,5]
Covid-19 (Centers for Disease Control [CDC]) case definition, defined as a positive post-baseline RT-PCR test result, together with eligible symptoms.
SARS-CoV-2 infection are defined as either a positive post-baseline bAb level against SARS-CoV-2 nucleocapsid protein or a positive post-baseline RT-PCR test in participants with negative SARS-CoV-2 status pre-booster.
Asymptomatic SARS-CoV-2 are identified by absence of symptoms in participants with infections as detected by RT-PCR or serology tests in participants with negative SARS-CoV-2 status pre-booster.
*Person-years is defined as the total time from randomization date to the date of SARS-CoV-2 infection, last date of study participation, or efficacy data cut-off date, whichever is the earliest.
†Incidence rate is defined as the number of participants with an event divided by the total person-years (total time at risk). The 95% CI is calculated using the exact method (Poisson distribution) and adjusted by person-years.

#### Table S9. Variant Sequences Detected in Covid-19 Cases, Part 2

| | mRNA-1273.214 (N=997) | mRNA-1273 (N=937) |
|---|---|---|
| Covid-19 cases ≥14 days post-randomization, n | 158 | 166 |
| with sequencing data, n (%)* | 135 (85.4) | 154 (92.8) |
| BA.2 and BA.2 Sublineages, n (%)† | 23 (17.0) | 32 (20.8) |
| BA.2 | 9 (6.7) | 15 (9.7) |
| BA.2 sublineages | 14 (10.4) | 17 (11.0) |
| BA.4 and BA.4 Sublineages, n (%)† | 18 (13.3) | 29 (18.8) |
| BA.4 | 10 (7.4) | 12 (7.8) |
| BA.4 sublineages n (%) | 8 (5.9) | 17 (11.0) |
| BA.5 and BA.5 Sublineages, n (%)† | 94 (69.6) | 93 (60.4) |
| BA.5 | 1 (0.7) | 5 (3.2) |
| BA.5 sublineages | 93 (68.9) | 88 (57.1) |

Percentages of variant sequence and sublineages detected by RT-PCR in nasopharyngeal swabs of participants positive for Covid-19 defined per the primary COVE definition[4,5] ≥14 days after randomization through the data cutoff date in the per-protocol efficacy set of part 2.
*Percentage based on all Covid-19 cases
†Percentages based on all Covid-19 cases with sequencing data.

**Table S10. Exploratory Analysis of Omicron Sublineage-Specific Covid-19 Cases Based on Primary Case Definition Starting 14 Days After Vaccine Booster - Per Protocol Efficacy Set, Part 2**

| | **mRNA-1273.214** (N=997) | **mRNA-1273** (N=937) |
|---|---|---|
| Overall Covid-19 cases (%) with or without sequencing data | 158 (15.8) | 166 (17.7) |
| Relative VE based on hazard ratio (95% CI)* | 11.4% (-10.2−28.7%) | |
| Covid-19 cases (%) with sequencing data | 135 (14.4) | 154 (16.4) |
| Number (%) of Covid-19 cases with BA.2/BA.2 sublineages | 23 (2.3) | 32 (3.4) |
| Number of Covid-19 cases with competing event | 135 | 134 |
| Relative VE based on hazard ratio (95% CI)† | 32.6% (-15.1−60.5%) | |
| Number (%) of Covid-19 cases with BA.4/BA.4 sublineages | 18 (1.8) | 29 (3.1) |
| Number of Covid-19 cases with competing event | 140 | 137 |
| Relative VE based on hazard ratio (95% CI)† | 41.6 % (-5.1−67.5%) | |
| Number (%) of Covid-19 cases with BA.5/BA.5 sublineages | 94 (9.4) | 93 (9.9) |
| Number of Covid-19 cases with competing event | 64 | 73 |
| Relative VE based on hazard ratio (95% CI)† | 4.4 % (-27.2−28.2%) | |
| Number (%) of Covid-19 cases with non-BA.5 sublineages (BA.2/BA.2 sublineages/BA.4/BA.4 sublineages) | 41 (4.1) | 61 (6.5) |
| Number of Covid-19 cases with competing event | 117 | 105 |
| Relative VE based on hazard ratio (95% CI)† | 37.3% (6.9−57.8%) | |

VE=vaccine efficacy. Percentages are based on participants in the per protocol set for efficacy in part 2. *The relative VE was estimated by a proportional hazards model in the mRNA-1273.214 versus mRNA-1273 group. †A competing risk method was used to analyze sublineage-specific events, where competing events were not censored. The Fine-Gray proportional hazards model for the subdistribution of a competing risk was used to estimate the hazard ratio and relative VE (1- hazard ratio) for the sublineages.[6]
